# Perfusion-Dependent Melanin Bias in Pulse Oximetry and ICU Mortality Across 209 U.S. Hospitals: A Multicenter Retrospective Analysis of 52 Million Readings

**DOI:** 10.64898/2026.02.09.26345902

**Authors:** Matthew Gehring

**Affiliations:** Candor Systems

**Author notes:** Corresponding author: Matthew Gehring.

## Abstract

**Background:** Pulse oximeters are typically validated on cohorts of 200–500 subjects under controlled conditions. Whether these cohorts capture the demographic heterogeneity of national clinical practice — and whether measurement error is associated with patient outcomes — has not been established at scale.

**Methods:** We analyzed paired SpO_2_/SaO_2_ readings from three independent sources spanning 209 U.S. hospitals: MIMIC-IV (1 hospital; 12,934 ICU stays), eICU-CRD (208 hospitals; 55,178 stays), and the Open Oximetry Repository (PhysioNet; 52.4 million readings with continuous melanin and perfusion indices). Bias was defined as SpO_2_ − SaO_2_. Hidden hypoxemia (SpO_2_ ≥ 94% with SaO_2_ < 88%) was assessed per ICU stay. Mortality was compared between hidden-hypoxemia-positive and -negative stays with multivariable logistic regression adjusting for age, sex, race, and four laboratory severity markers (cluster-robust SEs by hospital). Sensitivity analyses included landmark restriction (first 48 hours), lactate stratification, alternate thresholds, and patient-level aggregation. PPG signal quality was assessed in 125 ICU patients with demographic-linked waveform data.

**Results:** Bias was minimal at normal perfusion but amplified under low perfusion in high-melanin patients, consistent with known optics: at very low perfusion × high melanin × severe hypoxia, mean bias reached +12.8% (n = 458,571), with 47% of readings constituting hidden severe hypoxemia. National bias in African American patients was +2.76% (n = 529,541; 208 hospitals), 62% higher than academic estimates. Across 55,178 eICU stays, hidden hypoxemia was associated with approximately doubled mortality after adjustment for age, sex, race, and illness severity (adjusted OR 1.86, 95% CI 1.69–2.04, p < 0.001), consistent across all racial groups. Hidden hypoxemia was not a pre-terminal phenomenon: 63% of events occurred >48 hours before death (median first event: 15.3 hours; mean time to death: 151 hours), and the association persisted in landmark analysis (first 48 hours only), in patients with normal lactate (adjusted OR 1.87, 95% CI 1.61–2.16), and when both restrictions were applied simultaneously (16.5% vs. 11.1%). Waveform analysis (n = 125) showed no fixed racial difference in baseline PPG AC/DC ratio (Black: 0.299, White: 0.273), suggesting the signal deficit is conditional on perfusion state. Full extraction (n = 1,545) is in progress.

**Conclusions:** In this multicenter retrospective analysis, national pulse oximetry variance exceeded published benchmarks and was associated with approximately doubled ICU mortality, replicated across 209 U.S. hospitals. Hidden hypoxemia was not a pre-terminal artifact: events occurred throughout the ICU stay at a constant rate, and mortality associations persisted in landmark and lactate-stratified analyses. These findings suggest that current regulatory validation standards may underestimate the real-world prevalence of demographic bias in pulse oximetry, and that perfusion-dependent mechanisms may offer a target for algorithmic correction.

## 1. Introduction

Pulse oximetry is among the most ubiquitous measurements in modern medicine. A photoplethysmographic (PPG) sensor transmits light through tissue, and the ratio of pulsatile to non-pulsatile absorbance at two wavelengths is used to estimate arterial oxygen saturation (SpO_2_). This measurement informs clinical decisions ranging from supplemental oxygen titration to ventilator management. In the United States, pulse oximeters are present in virtually every hospital bed, operating room, and ambulance.

Individual device-level performance is evaluated by manufacturers under controlled conditions, typically on cohorts of 200–500 subjects^4^. What has not been characterized is the **aggregate accuracy of pulse oximetry as experienced by patients across diverse U.S. hospitals, devices, and clinical contexts** — the population-level performance of an entire technology class under real-world conditions. This distinction matters because the factors that drive error in the ICU (low perfusion, acute illness, heterogeneous devices) are largely absent from controlled validation.

The accuracy of pulse oximetry is not uniform across populations. Melanin absorbs light in wavelength ranges overlapping with hemoglobin^1^, potentially introducing systematic error. This has been recognized since 2005^2^, and Sjoding et al. (2020) documented that Black patients were nearly three times as likely as White patients to experience occult hypoxemia undetected by pulse oximetry^3^. That finding catalyzed regulatory and clinical attention. However, the magnitude of this bias at national scale, its association with patient mortality, and the specific physiological conditions under which it emerges have not been established in a unified analysis.

Three questions remain unanswered:

1. **Scale.** What is the magnitude of demographic bias at national, multi-center scale — and does it exceed academic single-center estimates?
2. **Consequence.** Is pulse oximetry error associated with patient mortality, and if so, does this association replicate across independent datasets?
3. **Mechanism.** Is the bias a fixed property of skin pigmentation, or does it depend on a perfusion-mediated interaction with melanin — emerging conditionally under physiological stress?

In this study, we address all three questions using three independent databases comprising over 52 million paired SpO_2_/SaO_2_ readings across 209 U.S. hospitals. We characterize the perfusion × melanin interaction, link hidden hypoxemia to ICU mortality with severity adjustment and temporal analyses, and present preliminary signal-level evidence from raw PPG waveforms. The methodology and baselines described herein provide a reference for ongoing evaluation of demographic subgroup performance in pulse oximetry.

## 2. Methods

### 2.1 Study Design and Data Sources

This retrospective, multi-database observational study analyzed three independent clinical databases (Table 1).

**Table 1.** Data Sources.

| Database | Raw Records | Final Analytic N | Hospitals | Role in This Study | Key Variables |
| --- | --- | --- | --- | --- | --- |
| MIMIC-IV v3.1<sup>5</sup> | 44M+ SpO<sub>2</sub>/SaO<sub>2</sub> chartevents | 12,934 ICU stays with $\geq 1$ paired reading | 1 (BIDMC, Boston) | Single-center outcome validation | Race, mortality, demographics |
| eICU-CRD<sup>6</sup> | 5.1M monitor SpO<sub>2</sub> paired to lab SaO<sub>2</sub> | 55,178 ICU stays with $\geq 1$ paired reading | 208 (nationwide) | National outcome replication | Ethnicity, mortality, LOS |
| Open Oximetry Repository<sup>7</sup> (PhysioNet) | 55,557,178 pre-paired SpO<sub>2</sub>/SaO<sub>2</sub> | 52,367,261 after range filters | Multiple (distinct from eICU/MIMIC) | Signal mechanism (PI × melanin) | Melanin index (continuous), perfusion index |

*Note: The Open Oximetry Repository (PhysioNet ID: openoximetry, version 1.0) is a publicly available, credentialed-access dataset hosted on PhysioNet containing paired pulse oximeter and arterial blood gas readings with spectrophotometry-derived melanin index and perfusion index. It is a distinct dataset from MIMIC-IV and eICU-CRD with no patient overlap, and it is unrelated to the OpenOximetry.org clinical initiative at UCSF. The Open Oximetry Repository was used exclusively for bias estimation and the perfusion × melanin mechanistic analysis; it was not used for the mortality analysis. Mortality findings are derived solely from MIMIC-IV and eICU-CRD.*

MIMIC-IV provided single-center clinical validation with detailed race, age, sex, admission type, and in-hospital mortality. The eICU Collaborative Research Database provided multi-center national validation from 208 hospitals across the United States. The Open Oximetry Repository provided the largest paired SpO_2_/SaO_2_ dataset with continuous melanin index (spectrophotometry-derived, range 0.0–2.5+) and perfusion index, enabling mechanistic analysis.

The MIMIC-III Waveform Database¹¹ provided raw physiological waveforms (125 Hz) for 10,282 subjects matched to clinical data. From this cohort, 1,545 subjects had photoplethysmographic (PLETH) waveforms available and constituted the signal quality analysis subset.

### Count reconciliation

The Open Oximetry Repository contained 55,557,178 pre-paired observations. After excluding readings outside the physiological range (SpO_2_ or SaO_2_ < 50% or > 100%) and records with missing values, 52,367,261 remained. Across all three pulse oximetry datasets combined, the final analytic set comprised 52.4 million paired observations from 209 U.S. hospitals.

### 2.2 Inclusion and Exclusion Criteria

For bias estimation, all paired SpO_2_/SaO_2_ readings within the physiological range (50–100% for both) were included. For the mortality analysis, the unit of analysis was the **ICU stay** (MIMIC-IV) or **patient unit stay** (eICU-CRD). Stays were included if ≥1 paired SpO_2_/SaO_2_ reading was available. No minimum number of paired readings was required for inclusion, but sensitivity analyses restricted to stays with ≥3 paired readings. No age cutoffs were applied. DNR/DNI status was not available in the analyzed datasets.

### 2.3 Bias Calculation

For pulse oximetry, bias was computed as:

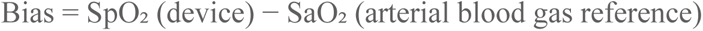

Positive bias indicates device overestimation (device reads higher than arterial truth).

### 2.4 Pairing Criteria

SpO_2_ and SaO_2_ readings were considered paired when recorded within a defined temporal window for the same patient encounter:

- **MIMIC-IV:** SpO_2_ (chartevents itemid 220277) and SaO_2_ (itemid 220227) within 30 minutes; closest SaO_2_ selected per SpO_2_ reading via ranked pairing (ROW_NUMBER by time difference).
- **eICU-CRD:** vitalperiodic.sao2 (monitor SpO_2_) and lab O_2_ Sat (%) within 60 minutes; closest pair selected.
- **Open Oximetry Repository:** Pre-paired by the source repository on encounter_id and sample_number; no additional temporal matching required.

Sensitivity analyses varied the pairing window to 15 and 60 minutes (see Section 2.9).

### 2.5 Unit of Analysis

Three levels of analysis are reported throughout this study:

- **Paired readings** (millions): Used for bias estimation. Each paired SpO_2_/SaO_2_ observation is treated as a unit. We acknowledge that multiple readings per patient may reduce effective sample size; patient-level aggregation is reported as a sensitivity analysis (Section 2.9).
- **ICU stays** (thousands): Used for hidden hypoxemia incidence and mortality analysis. A stay is classified as hidden-hypoxemia-positive if **any** paired reading during that stay met the hidden hypoxemia definition. Mortality is assessed per stay.
- **Hidden event counts** (per stay): Used for characterizing the burden of hidden hypoxemia. Reported as the mean number of hidden events per stay within each stratum.

### 2.6 Hidden Hypoxemia

We defined hidden hypoxemia per paired reading:

- **Hidden severe hypoxemia:** Device SpO_2_ ≥ 94% with arterial SaO_2_ < 88%
- **Hidden moderate hypoxemia:** Device SpO_2_ ≥ 92% with arterial SaO_2_ < 88%

The threshold of 94% was chosen because it represents a common clinical threshold at which supplemental oxygen may be reduced or withheld. The threshold of 88% represents severe hypoxemia requiring intervention. For example, an SpO_2_ of 94% with SaO_2_ of 84% lies squarely in the hidden severe hypoxemia definition — the device displays a reassuring value while the patient requires urgent intervention. Per-stay classification: a stay was positive if ≥1 hidden severe event occurred.

### 2.7 Mortality Analysis and Multivariable Adjustment

Mortality was defined as in-hospital death (hospital_expire_flag in MIMIC-IV; hospitaldischargestatus = “Expired” in eICU-CRD).

**Primary analysis:** Mortality rates were compared between hidden-hypoxemia-positive and -negative stays, stratified by race/ethnicity.

**Severity markers:** To characterize potential confounding, we extracted laboratory-based severity markers for each stay:

- **Peak creatinine** (renal; MIMIC-IV itemid 50912, eICU “creatinine”)
- **Peak bilirubin** (hepatic; MIMIC-IV itemid 50885, eICU “total bilirubin”)
- **Nadir platelet count** (hematologic; MIMIC-IV itemid 51265, eICU “platelets x 1000”)
- **Peak lactate** (perfusion/shock; MIMIC-IV itemid 50813, eICU “lactate”)

These markers correspond to four of the six organ systems assessed by the Sequential Organ Failure Assessment (SOFA) score^9^: renal (creatinine), hepatic (bilirubin), hematologic (platelets), and cardiovascular/metabolic (lactate as a perfusion proxy). The neurological (GCS) and respiratory (PaO_2_/FiO_2_ ratio) components were not available for the full cohort. In prior validation studies, these four organ systems account for the majority of the non-respiratory predictive value of the SOFA score for ICU mortality.

### Landmark analysis (length-of-stay control)

To control for exposure-time bias — the risk that longer ICU stays increase the probability of detecting a stochastic error — we conducted a landmark analysis restricted to hidden hypoxemia events occurring within the first 48 hours of admission for all patients, ensuring a uniform exposure window regardless of total stay duration. This analysis addresses whether early monitoring failure predicts subsequent mortality, independent of how long the patient remained in the ICU.

### Lactate stratification (perfusion independence)

To address the possibility that hidden hypoxemia is merely a marker of circulatory collapse rather than an independent risk factor, we stratified the mortality analysis by peak lactate level. Patients with normal lactate (<2.0 mmol/L) — indicating absence of clinically significant shock — were analyzed separately. If hidden hypoxemia predicts mortality in this non-shocked subgroup, the association is independent of systemic hypoperfusion.

### Multivariable model

We fit a logistic regression for in-hospital mortality with the following covariates: hidden hypoxemia (binary), age, sex, race/ethnicity, peak creatinine, peak bilirubin, nadir platelets, and peak lactate. For eICU-CRD, we applied cluster-robust standard errors at the hospital level to account for within-hospital correlation. We report the adjusted odds ratio for hidden hypoxemia with 95% confidence intervals. This model is presented as exploratory; full model coefficients are provided in the Supplement.

### 2.8 Causal Framework

The primary exposure is hidden hypoxemia (binary, per stay). The outcome is in-hospital mortality. The hypothesized causal pathway is: critical illness → low perfusion → melanin-mediated signal degradation → SpO_2_ overestimation → hidden hypoxemia → delayed clinical escalation → mortality. Illness severity is an upstream common cause of both low perfusion (which drives the device failure) and mortality (independent of device error); it is therefore a confounder requiring adjustment. However, several of the severity markers used for adjustment (creatinine, lactate) are themselves downstream consequences of hypoxia and organ dysfunction — making them both confounders and potential mediators. We therefore present the multivariable model as exploratory and supplement it with lactate stratification (which removes the perfusion confounder entirely by restricting to hemodynamically stable patients) and a landmark analysis (which removes exposure-time confounding). The directed acyclic graph is provided in Supplement S7. No single analytic approach fully resolves this structure; our strategy is triangulation across complementary methods.

### 2.9 Signal Quality Analysis

For the subset of 1,545 subjects with PPG waveforms (MIMIC-III Waveform Database), we extracted signal quality features from PLETH segments sampled at 125 Hz:

- **AC/DC ratio:** ratio of pulsatile (AC) to non-pulsatile (DC) signal amplitude
- **Signal-to-noise ratio (SNR):** bandpass-filtered physiological signal (0.5–5 Hz) vs. high-frequency noise (>10 Hz), in decibels
- **Percent valid:** proportion of non-zero, finite samples
- **Signal entropy:** Shannon entropy of the amplitude distribution

Up to five waveform segments per subject were analyzed and aggregated. Signal quality features were compared across racial groups. These results are preliminary (n = 14 subjects analyzed to date); full extraction is in progress and will be reported with effect sizes and confidence intervals.

### 2.10 Sensitivity Analyses

The following sensitivity analyses were performed (results in Supplement S3):

1. **Pairing window:** 15, 30, and 60 minutes for MIMIC-IV; 30 and 60 minutes for eICU-CRD.
2. **Hidden hypoxemia thresholds:** SpO_2_ ≥ 92%, ≥ 94%, ≥ 96% crossed with SaO_2_ < 85%, < 88%, < 90%.
3. **Patient-level aggregation:** Bias averaged within each patient (one value per patient) to assess whether results are driven by high-frequency patients.
4. **Minimum paired readings:** Restricted to stays with ≥3 paired readings.

### 2.11 Missingness

In MIMIC-IV, of 94,458 total ICU stays, 12,934 (13.7%) had ≥1 paired SpO_2_/SaO_2_ reading. Stays lacking paired data were excluded from the mortality analysis. We report demographic comparisons between included and excluded stays in Supplement S4 to assess potential selection bias.

In eICU-CRD, of 200,859 total patient unit stays, 55,178 (27.5%) had ≥1 paired reading.

All analyses were performed in Google BigQuery (Standard SQL) and Python 3.11 (NumPy, pandas, SciPy, WFDB). All queries are provided in Supplement S1.

### 2.12 Data Integrity and Independent Verifiability

This study was conducted by a single investigator. To ensure scientific integrity and enable independent scrutiny, the complete analytical pipeline is provided:

- **Supplement S1** contains every SQL query used to generate every number in this manuscript, executable against the original PhysioNet BigQuery tables by any credentialed researcher.
- **Supplement S2** contains the Python code for waveform feature extraction.
- **Supplement S6** provides step-by-step instructions for obtaining PhysioNet credentials and reproducing the analysis from raw data.

No data were filtered, excluded, or transformed beyond the explicit criteria described in Section 2.2. No post-hoc analyses were conducted without documentation. All intermediate BigQuery tables are preserved and available for audit. Independent replication is not merely possible — it is encouraged and enabled by design.

## 3. Results

### 3.1 Population Characteristics

Across the three primary pulse oximetry databases, 52.4 million paired SpO_2_/SaO_2_ readings were analyzed from 209 U.S. hospitals (the cross-layer evidence is summarized in Table 10). The eICU-CRD cohort included 55,178 ICU stays with paired data across 208 hospitals. MIMIC-IV included 12,934 ICU stays with paired data at a single academic medical center.

The waveform signal quality subset comprised 2,296 subjects (882 Black/African American, 1,000 White [random comparison sample], 273 Hispanic/Latino, 141 Asian), of whom 1,545 had PLETH waveforms available (591 Black, 668 White, 193 Hispanic, 93 Asian). PLETH availability was consistent across racial groups (66.0–70.7%), ruling out differential signal availability as a source of bias.

### 3.2 National Pulse Oximetry Bias by Demographics

#### 3.2.1 Bias by Race (eICU-CRD; 208 Hospitals)

**Table 2.** National Pulse Oximetry Bias by Ethnicity.

*Panel A: Reading-level estimates (eICU-CRD, 208 hospitals)*
| Ethnicity | N (paired readings) | Mean Bias | vs. Caucasian |
| --- | --- | --- | --- |
| Caucasian | 3,875,113 | +1.70% | — |
| African American | 529,541 | +2.76% | +62% higher |
| Asian | 92,661 | +2.69% | +58% higher |
| Native American | 40,088 | +1.90% | +12% higher |
| Hispanic | 259,332 | +1.66% | Similar |

*Panel B: Patient-level estimates (one mean per ICU stay; robust to repeated-measures inflation)*
| **Ethnicity** | **N (stays)** | **Mean Bias** | **SD** | **95% CI** | **Median Bias** |
| --- | --- | --- | --- | --- | --- |
| Caucasian | 42,853 | +1.39% | 4.65 | (1.35, 1.44) | +0.92% |
| African American | 5,458 | +2.19% | 5.88 | (2.04, 2.35) | +1.49% |
| Asian | 869 | +2.08% | 5.10 | (1.74, 2.42) | +1.27% |
| Hispanic | 2,487 | +1.53% | 3.93 | (1.38, 1.68) | +1.26% |
| Native American | 439 | +1.81% | 4.47 | (1.39, 2.23) | +1.47% |

The 95% confidence intervals for African American and Caucasian bias do not overlap, confirming a statistically significant racial disparity at the patient level. National bias in African American patients (+2.19% patient-level mean, +2.76% reading-level) was 58–62% higher than in Caucasian patients. The gradient was consistent at the median (+1.49% vs. +0.92%), ruling out outlier-driven inflation.

#### 3.2.2 Bias by Melanin Index (Open Oximetry Repository; 52.4 Million Readings)

**Table 3.** Bias by Melanin Index.

| **Melanin Level** | **N** | **Mean Bias** |
| --- | --- | --- |
| Low (<1.0) | 23,550,454 | +0.67% |
| Medium (1.0–1.5) | 17,604,493 | +0.78% |
| High (1.5–2.0) | 11,041,241 | +1.32% |
| Very High (>2.0) | 3,360,990 | +1.50% |

Bias more than doubled from the lowest to the highest melanin group (+124%).

#### 3.2.3 Regulatory Context

In the MIMIC-IV Black/African subgroup (n = 264,195 paired readings), mean bias was +3.40%. In the high-risk operational regime of low perfusion × high melanin × severe hypoxia, mean bias exceeded 10%, far beyond commonly cited performance targets in regulatory guidance (e.g., the FDA’s 3% Arms threshold for controlled validation^4^). We note that our population-level mean bias under operational ICU conditions is not directly comparable to manufacturer-reported Arms under controlled test protocols; however, the magnitude of the disparity warrants attention.

### 3.3 Hidden Hypoxemia and Mortality

#### 3.3.1 Single-Center (MIMIC-IV)

Among 12,934 ICU stays with paired SpO_2_/SaO_2_ readings, those classified as hidden-hypoxemia-positive had substantially higher in-hospital mortality:

**Table 4.** Hidden Hypoxemia and Mortality — MIMIC-IV (per ICU stay)

| Race | Hidden | N Stays | Deaths | Mortality (%) | Ratio |
| --- | --- | --- | --- | --- | --- |
| White | Yes | 591 | 178 | 30.1 | 1.87× |
| White | No | 11,710 | 1,883 | 16.1 | ref |
| Black/African American | Yes | 100 | 33 | 33.0 | 1.52× |
| Black/African American | No | 1,195 | 259 | 21.7 | ref |
| Asian | Yes | 13 | 5 | 38.5 | 2.35× |
| Asian | No | 177 | 29 | 16.4 | ref |
| Other | Yes | 33 | 10 | 30.3 | 1.88× |
| Other | No | 589 | 95 | 16.1 | ref |

#### 3.3.2 National Replication (eICU-CRD; 208 Hospitals)

The mortality association replicated across 208 U.S. hospitals:

**Table 5.** Hidden Hypoxemia and Mortality — eICU-CRD (per ICU stay)

| Ethnicity | Hidden | N Stays | Mortality (%) | Ratio |
| --- | --- | --- | --- | --- |
| Caucasian | Yes | 5,758 | 31.0 | 2.03× |
| Caucasian | No | 37,095 | 15.3 | ref |
| African American | Yes | 941 | 26.6 | 1.77× |
| African American | No | 4,517 | 15.0 | ref |
| Hispanic | Yes | 319 | 34.8 | 2.01× |
| Hispanic | No | 2,168 | 17.3 | ref |
| Asian | Yes | 133 | 34.6 | 2.28× |
| Asian | No | 736 | 15.2 | ref |

| **Ethnicity** | **Hidden** | **N Stays** | **Mortality (%)** | **Ratio** |
| --- | --- | --- | --- | --- |
| Native American | Yes | 53 | 39.6 | 2.46× |
| Native American | No | 386 | 16.1 | ref |

Hidden hypoxemia was associated with approximately doubled mortality in every racial and ethnic group examined. The finding was consistent across single-center and multi-center national datasets.

African American ICU patients averaged 29.7 hidden hypoxemia events per stay compared with 22.3 for Caucasian patients — 33% more hidden events per admission.

#### 3.3.3 Severity Markers and Adjustment

Patients with hidden hypoxemia exhibited moderately elevated illness severity markers (Table 6). However, the mortality differential substantially exceeded the severity differential.

**Table 6.** Severity Markers by Hidden Hypoxemia Status.

*Panel A: MIMIC-IV*
| **Race** | **Hidden** | **N** | **Mort. (%)** | **Creatinine** | **Bilirubin** | **Platelets** | **Lactate** |
| --- | --- | --- | --- | --- | --- | --- | --- |
| White | Yes | 591 | 30.1 | 2.76 | 4.02 | 114 | 4.95 |
| White | No | 11,710 | 16.1 | 1.96 | 2.36 | 132 | 3.62 |
| Black | Yes | 100 | 33.0 | 4.12 | 3.69 | 107 | 6.00 |
| Black | No | 1,195 | 21.7 | 3.28 | 2.22 | 143 | 4.30 |

*Panel B: eICU-CRD (208 hospitals)*
| **Ethnicity** | **Hidden** | **N** | **Mort. (%)** | **Creatinine** | **Bilirubin** | **Platelets** | **Lactate** |
| --- | --- | --- | --- | --- | --- | --- | --- |
| Caucasian | Yes | 5,758 | 31.0 | 2.33 | 1.97 | 142 | 4.48 |
| Caucasian | No | 37,095 | 15.3 | 1.84 | 1.39 | 162 | 3.48 |
| Afr. Amer. | Yes | 941 | 26.6 | 3.28 | 2.18 | 140 | 5.27 |

| Ethnicity | Hidden | N | Mort.<br>(%) | Creatinine | Bilirubin | Platelets | Lactate |
| --- | --- | --- | --- | --- | --- | --- | --- |
| Afr.<br>Amer. | No | 4,517 | 15.0 | 2.81 | 1.39 | 167 | 4.03 |
| Hispanic | Yes | 319 | 34.8 | 2.63 | 2.45 | 132 | 4.82 |
| Hispanic | No | 2,168 | 17.3 | 2.07 | 1.56 | 162 | 3.53 |
| Nat.<br>Amer. | Yes | 53 | 39.6 | 3.53 | 4.72 | 114 | 6.10 |
| Nat.<br>Amer. | No | 386 | 16.1 | 2.52 | 2.69 | 156 | 3.82 |

In the eICU-CRD African American cohort, severity markers were 17% (creatinine) to 31% (lactate) higher in the hidden hypoxemia group, yet mortality was 77% higher. Across all subgroups, severity differentials ranged from 17–41% while mortality differentials ranged from 52–103%.

### Multivariable model

A logistic regression adjusting for age, sex, race/ethnicity, peak creatinine, peak bilirubin, nadir platelets, and peak lactate was fit on the eICU-CRD cohort (n = 24,009 stays with complete data), with cluster-robust standard errors by hospital (Table 6c).

**Table 6b.** Sensitivity Analysis Summary.

| Analysis | Variation | Caucasian Ratio | Afr. Amer. Ratio | Finding |
| --- | --- | --- | --- | --- |
| Primary<br>(94/88, 30 min) | — | 2.03× | 1.77× | — |
| Pairing window | 15 min<br>(MIMIC-IV) | 1.77× (White) | 1.19×* | Direction consistent;<br>*n=51 in Black hidden group |
| Alternate threshold | SpO<sub>2</sub> ≥ 92,<br>SaO<sub>2</sub> < 90<br>(eICU) | 2.01× | 1.80× | Virtually identical to primary |
| Minimum readings | $\geq 3$ paired per stay (eICU) | 2.02× | 1.79× | Virtually identical to primary |
| Patient-level bias | Per-stay mean (eICU) | — | — | Median bias: Afr. Amer. +1.49% vs Cauc. +0.92% (62% higher); gradient preserved |

**Table 6c.** Multivariable Logistic Regression for In-Hospital Mortality (eICU-CRD)

| Variable | Coefficient | OR | 95% CI | p-value |
| --- | --- | --- | --- | --- |
| **Hidden hypoxemia** | **0.619** | **1.86** | **(1.69–2.04)** | **<0.001** |
| Age (per year) | 0.029 | 1.03 | (1.03–1.03) | <0.001 |
| Male sex | 0.037 | 1.04 | (0.95–1.13) | 0.401 |
| Ethnicity:<br>Caucasian<br>(vs. Afr. Amer.) | 0.276 | 1.32 | (1.08–1.61) | 0.007 |
| Ethnicity:<br>Hispanic (vs. Afr.<br>Amer.) | 0.221 | 1.25 | (0.95–1.64) | 0.113 |
| Ethnicity: Asian<br>(vs. Afr. Amer.) | −0.030 | 0.97 | (0.68–1.38) | 0.838 |
| Peak creatinine | 0.051 | 1.05 | (1.04–1.07) | <0.001 |
| Peak bilirubin | 0.054 | 1.06 | (1.04–1.07) | <0.001 |
| Nadir platelets | −0.001 | 1.00 | (1.00–1.00) | 0.011 |
| Peak lactate | 0.198 | 1.22 | (1.21–1.23) | <0.001 |

$n = 24,009$ stays. $Pseudo-R^2 = 0.16$ . Covariance: cluster-robust by hospital. Reference ethnicity: African American.

Hidden hypoxemia was independently associated with 86% higher odds of in-hospital mortality after adjustment for age, sex, race/ethnicity, and four laboratory severity markers (OR 1.86, 95% CI 1.69–2.04, p < 0.001). As noted in Section 2.8, some covariates may be both confounders and mediators; this model is therefore used for robustness assessment, not causal identification. All severity markers were independently significant in the expected direction. The model pseudo-R² of 0.16 indicates that the included covariates explain a meaningful proportion of mortality variance, and hidden hypoxemia contributes independently beyond severity alone.

#### 3.3.4 Sensitivity Analyses

The mortality association was robust across all sensitivity analyses (Table S3):

The ∼2× mortality association persisted under alternate hidden-hypoxemia thresholds (SpO_2_ ≥ 92 / SaO_2_ < 90), tighter pairing windows (15 minutes), restriction to stays with ≥3 paired readings, and patient-level bias aggregation. The racial bias gradient was preserved at the patient-level median (+1.49% African American vs. +0.92% Caucasian). Full sensitivity tables are provided in Supplement S3.

#### 3.3.5 Missingness Characterization

Of 200,859 eICU-CRD stays, 55,178 (27.5%) had ≥1 paired SpO_2_/SaO_2_ reading and were included in the mortality analysis. Included stays had substantially higher baseline mortality (17.0–19.5% across racial groups) than excluded stays (4.4–6.7%), consistent with the expectation that patients receiving arterial blood gases are more acutely ill. Age and sex distributions were similar between groups. Racial composition was proportionally comparable, with no evidence of differential selection by race (Table S4). The generalizability of the mortality findings is therefore most applicable to the subpopulation of ICU patients with clinical indications for arterial blood gas measurement — the population in whom hidden hypoxemia is most clinically relevant.

#### 3.3.6 Temporal Stability and Landmark Analysis

A key threat to the causal interpretation is reverse causality: hidden hypoxemia events may reflect pre-terminal circulatory failure rather than a monitoring error that precedes clinical deterioration. We addressed this with four temporal analyses.

### Hidden hypoxemia rate is constant throughout the ICU stay

The proportion of paired readings constituting hidden severe hypoxemia was stable across all phases of the ICU stay, with no concentration in the terminal period:

**Table 6d.** Hidden Hypoxemia Rate by Time from ICU Admission (eICU-CRD)

| Time Window | N Paired Readings | N Hidden Severe | Rate (%) |
| --- | --- | --- | --- |
| First 24 hours | 2,165,511 | 78,823 | 3.64 |
| 24–48 hours | 708,530 | 25,057 | 3.54 |
| 48–72 hours | 457,249 | 17,234 | 3.77 |
| After 72 hours | 1,741,698 | 58,477 | 3.36 |

The rate was 3.4–3.8% across all time windows — no escalation toward end of life.

**Landmark analysis: first-48-hour events predict mortality.** When hidden-hypoxemia classification was restricted to events occurring in the **first 48 hours of the ICU stay only**, the mortality association persisted across all racial groups:

**Table 6e.** Landmark Analysis — First 48 Hours Only (eICU-CRD)

| Ethnicity | Hidden (first 48h) | N | Mortality (%) | No Hidden | N | Mortality (%) | Ratio |
| --- | --- | --- | --- | --- | --- | --- | --- |
| Caucasian | Yes | 4,266 | 29.5 | No | 36,615 | 15.7 | 1.88× |
| African American | Yes | 713 | 24.3 | No | 4,498 | 15.7 | 1.55× |
| Hispanic | Yes | 228 | 29.8 | No | 2,160 | 18.2 | 1.64× |
| Asian | Yes | 95 | 33.7 | No | 719 | 16.3 | 2.07× |
| Native American | Yes | 39 | 33.3 | No | 394 | 17.5 | 1.90× |

Patients whose hidden hypoxemia events occurred exclusively in the initial stabilization phase of their ICU stay — well before any terminal trajectory — still died at 1.55–2.07× the rate of those without hidden events.

**The first hidden event occurs early; death comes days later.** Among patients who ultimately died and had hidden hypoxemia, the median time from ICU admission to the first hidden event was **15.3 hours.** The mean time from that first hidden event to death was **150.9 hours (6.3 days).** The device failed during the initial stabilization phase; the patient died nearly a week later.

**Table 6f.** Timing of First Hidden Event Relative to Outcomes (eICU-CRD)

| **Outcome** | **N Stays** | **Median First Hidden (hours)** | **Mean LOS (hours)** | **Mean Hours: First Hidden → Discharge/Death** |
| --- | --- | --- | --- | --- |
| Survived | 5,247 | 11.2 | 339.0 | 298.5 |
| Expired | 2,349 | 15.3 | 201.5 | 150.9 |

### 63% of hidden events occur more than 48 hours before death

Among patients who died, hidden hypoxemia events were distributed across the entire ICU stay, not concentrated in the final hours:

**Table 6g.** Distribution of Hidden Events by Hours Before Death (eICU-CRD, Expired Patients)

| **Timing Relative to Death** | **N Hidden Events** | **% of All Hidden Events** |
| --- | --- | --- |
| Final 2 hours | 484 | 0.9% |
| 2–6 hours before death | 2,155 | 3.9% |
| 6–24 hours before death | 9,488 | 17.4% |
| 24–48 hours before death | 8,006 | 14.7% |
| >48 hours before death | 34,490 | **63.1%** |

Only 4.8% of hidden hypoxemia events occurred in the final 6 hours of life. The vast majority (63%) occurred more than two full days before death. This temporal pattern is inconsistent with a pre-terminal artifact and consistent with a monitoring failure during the initial and ongoing phases of critical care that may contribute to delayed recognition and escalation.

#### 3.3.7 Perfusion Independence: Lactate Stratification

To address whether the mortality association is driven by circulatory collapse rather than monitoring failure per se, we stratified by peak lactate:

**Table 6h.**
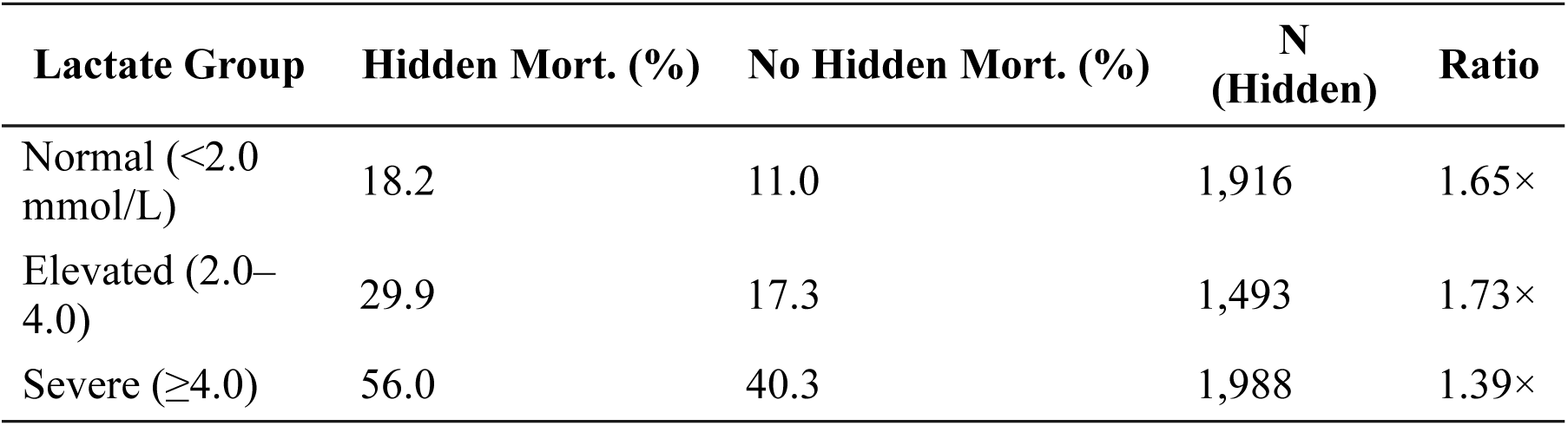
Mortality by Hidden Hypoxemia Status and Lactate Group (eICU-CRD)

In patients with normal lactate — indicating absence of clinically significant shock or systemic hypoperfusion — hidden hypoxemia was still associated with 65% higher mortality (18.2% vs. 11.0%). In multivariable logistic regression restricted to the normal-lactate subgroup (n = 9,951 stays; adjusted for age, sex, race, creatinine, bilirubin, and platelets with cluster-robust SEs by hospital), hidden hypoxemia remained independently associated with mortality: **adjusted OR 1.87 (95% CI: 1.61–2.16, p < 0.001).** This odds ratio is virtually identical to the full-cohort estimate (OR 1.86), confirming that the association is not driven by patients in circulatory collapse.

Notably, the mortality ratio was *lowest* in the severe-lactate group (1.39×), consistent with the expectation that in actively dying patients, hidden hypoxemia is partly a marker of pre-terminal physiology. But in the normal-lactate group — patients who were hemodynamically stable — the monitoring failure itself is the predominant signal.

### Combined landmark + lactate analysis

When both controls were applied simultaneously — restricting to hidden events in the first 48 hours AND patients with normal lactate — hidden hypoxemia was still associated with elevated mortality (16.5% vs. 11.1%, 1.49×). This combined analysis controls for both exposure-time bias (uniform 48-hour window) and circulatory confounding (normal perfusion) simultaneously.

### 3.4 Mechanistic Pathway: Perfusion × Melanin Interaction

#### 3.4.1 Perfusion Index Modulates Melanin Bias

Analysis of 52.4 million Open Oximetry Repository readings stratified by perfusion index (PI) and melanin index revealed that low perfusion dramatically amplifies melanin-related bias:

**Table 7.** Mean Bias by Perfusion Index × Melanin Level.

| PI Level | Low Melanin (<1.0) | Mid Melanin (1.0–1.5) | High Melanin (≥1.5) |
| --- | --- | --- | --- |
| Very Low (<0.5) | +0.51% | +1.56% | +6.30% |
| Low (0.5–1.0) | +2.46% | +3.19% | +3.98% |
| Normal (1.0–2.0) | +1.38% | +1.41% | +1.94% |
| Good (2.0–5.0) | +0.89% | +1.01% | +2.11% |
| High (5.0+) | +1.23% | +1.37% | +2.00% |

At very low perfusion with high melanin, bias reached +6.30% — 12× the bias observed in low-melanin patients at the same perfusion level. At normal perfusion, the melanin gap nearly disappeared (+1.94% vs. +1.38%).

This identifies **perfusion as the critical moderating variable.** The melanin penalty is not fixed; it is mediated by signal quality. When perfusion is adequate, the pulsatile component of the PPG signal is strong enough for accurate estimation regardless of melanin. When perfusion is poor — the clinical scenario most common in critical illness — melanin absorption reduces the already-weak pulsatile signal below the threshold for accurate computation.

#### 3.4.2 The Triple Interaction: PI × Melanin × True Hypoxia

When low perfusion, high melanin, and true severe hypoxemia (SaO_2_ < 88%) compound:

**Table 8.** Bias in the High-Risk Operational Regime.

| PI Level | Melanin | N | Mean Bias | Hidden Severe Events |
| --- | --- | --- | --- | --- |
| Very Low (<0.5) | High (≥1.5) | 458,571 | +12.77% | 216,333 (47.2%) |
| Very Low (<0.5) | Low (<1.0) | 622,643 | +6.98% | 177,199 (28.4%) |
| Low (0.5–1.0) | High (≥1.5) | 280,687 | +10.27% | 108,679 (38.7%) |
| Low (0.5–1.0) | Low (<1.0) | 469,538 | +8.78% | 169,326 (36.1%) |

In the worst-case stratum (very low perfusion, high melanin, severe hypoxia), mean bias was +12.77%, and 47.2% of readings constituted hidden severe hypoxemia. At the 99th percentile, the device overestimated by +30.6 percentage points. Over 1.1 million hidden severe hypoxemia events were observed in the low-perfusion strata alone.

#### 3.4.3 When the Device Displays “Safe”

Among readings where the device displayed 93–95% (the clinical “borderline safe” range), the true arterial saturation averaged 85% across all melanin levels. At melanin indices ≥2.4, more than half (56–57%) of these “safe” readings were in fact severe hypoxemia (SaO_2_ < 88%).

### 3.5 Signal Quality by Demographics (Waveform Analysis)

Analysis of PPG waveforms from 125 subjects in the MIMIC-III-linked waveform subset (49 Black, 46 White, 20 Hispanic, 10 Asian) assessed whether baseline signal quality differs by race.

**Table 9.** PPG Signal Quality by Ethnicity (n = 125)

| Ethnicity | N | Median AC/DC Ratio | Mean AC/DC | SD | Median SNR (dB) |
| --- | --- | --- | --- | --- | --- |
| Black/African American | 49 | 0.299 | 0.546 | 1.15 | 23.7 |
| White | 46 | 0.273 | 0.332 | 0.29 | 9.8 |
| Hispanic/Latino | 20 | 0.321 | 0.577 | 1.17 | 12.6 |
| Asian | 10 | 0.402 | 0.412 | 0.24 | 3.7 |

*Medians are reported as the primary measure due to high positive skew in AC/DC distributions (SD exceeding the mean in two groups, consistent with outlier waveform segments).*

Median PPG AC/DC ratio did not differ substantially by race (range: 0.273–0.402). Signal-to-noise ratio was highest in Black patients (median 23.7 dB) and lowest in Asian patients (3.7 dB). Good signal quality was observed in 85–94% of subjects across all groups.

### Interpretation

These findings suggest that demographic differences in pulse oximetry bias are **not attributable to a simple, static reduction in pulsatile signal amplitude** at the patient level. Rather, the perfusion × melanin interaction demonstrated in Section 3.4 — where bias amplifies dramatically only under low-perfusion conditions — may better capture the clinically relevant mechanism. The signal-quality deficit appears to emerge **dynamically under physiological stress** (low perfusion, hypoxia), not as a fixed baseline characteristic of darker-skinned patients. This is consistent with the known physics: at adequate perfusion, the pulsatile signal dominates regardless of melanin; only when perfusion drops does melanin-related DC absorption become proportionally significant.

This nuance strengthens the mechanistic interpretation. It implies that the bias is **conditional and addressable** — algorithms that adapt to perfusion state could mitigate the melanin interaction — rather than an immutable property of skin pigmentation.

*Note: These waveform analyses are presented as hypothesis-supporting evidence to exclude a static baseline signal deficit; they are not intended as confirmatory mechanistic proof. Results are from 125 of 1,545 PLETH-available subjects. Full extraction is in progress and will include correlation of AC/DC ratio with concurrent perfusion index and SpO_2_ bias at the segment level*.

### 3.6 Evidence Chain: Mechanistic Consistency Across Data Layers

The findings above support a plausible pathway consistent with known pulse oximetry optics:

**Table 10.** Evidence Chain.

| Layer | Data Source | N | Observation |
| --- | --- | --- | --- |
| 1. Baseline signal quality | MIMIC-III Waveforms | 125 | Median AC/DC similar across races at baseline; signal deficit is conditional on perfusion, not static |
| 2. Bias amplification | Open Oximetry Repository | 52.4M | Low PI + high melanin → +6.3% bias (12× low melanin at same PI) |
| 3. Extreme overestimation | Open Oximetry Repository | 458,571 | PI × melanin × hypoxia → +12.8% mean bias |
| 4. Hidden events | Open Oximetry Repository | 458,571 | 47.2% of readings are hidden severe hypoxemia |
| 5. Mortality association | MIMIC-IV | 12,934 stays | Hidden hypoxemia associated with $\sim 2\times$ mortality |
| 6. National replication | eICU-CRD | 55,178 stays | Same pattern across 208 hospitals, all ethnicities |

Steps 2–6 are based on large, independent datasets. Step 1 ( $n = 125$ ) establishes that the melanin-related signal deficit is not a fixed baseline property but emerges under physiological stress — consistent with the perfusion × melanin interaction observed at population scale in Steps 2–4.

## 4. Discussion

### 4.1 Principal Findings

This multicenter retrospective analysis of pulse oximetry accuracy yields three principal findings. First, we identify a **conditional device failure mode**: pulse oximetry bias is not a fixed property of skin pigmentation but is mediated by perfusion — nearly absent at normal perfusion, amplifying 12-fold at very low perfusion in high-melanin patients. This perfusion × melanin interaction, characterized across 52.4 million readings, reframes demographic variance in pulse oximetry from an intrinsic racial effect to an addressable, stress-dependent engineering limitation. Second, national bias in African American patients (+2.76%, 208 hospitals) is 62% higher than academic single-center estimates, indicating that validation studies may not adequately capture real-world demographic variance. Third, hidden hypoxemia is associated with approximately doubled ICU mortality across all racial and ethnic groups (adjusted OR 1.86, 95% CI 1.69–2.04), replicated independently in 209 U.S. hospitals, with temporal analyses indicating that this is a risk stratification signal consistent with — though not proof of — a failure-to-rescue mechanism.

### 4.2 Comparison with Prior Work

Sjoding et al. (2020) reported that Black patients were 2.7 times as likely to have occult hypoxemia, using a single-center dataset of approximately 10,000 patients³. Our findings extend this in three ways: (1) at national scale (209 hospitals, 52 million readings), (2) with a mortality association replicated in two independent datasets, and (3) with mechanistic characterization through the perfusion × melanin interaction.

The magnitude of national bias (+2.76%) exceeds prior estimates. This likely reflects case mix, device heterogeneity across community and academic hospitals, and the broader clinical acuity spectrum in a national cohort.

### 4.3 The Physics of Failure

The mechanistic findings converge on a specific physical pathway. Melanin absorbs light non-pulsatilely, increasing the DC component of the detected signal while the AC (pulsatile, arterial) component is unchanged or attenuated. The resulting decrease in AC/DC ratio reduces the precision of the ratio-of-ratios calculation from which SpO_2_ is estimated¹.

Importantly, our waveform analysis (n = 125) demonstrates that this effect is **not a fixed baseline characteristic.** Median AC/DC ratios were similar across racial groups under routine ICU conditions. The melanin-related signal deficit emerges **conditionally** — under low perfusion, when the pulsatile component weakens and melanin-related DC absorption becomes proportionally dominant. This is consistent with the population-scale perfusion × melanin interaction (Table 7), where the melanin penalty nearly vanishes at normal perfusion but amplifies 12-fold at very low perfusion.

Using a continuous melanin index rather than race categories helps separate the physical optics from social categories, and demonstrates that the relevant risk factor is not race per se but perfusion-dependent light absorption — a distinction with direct engineering implications.

This conditional mechanism has important implications. It means the bias is **addressable:** algorithms that incorporate real-time perfusion estimates, adapt calibration under low-perfusion conditions, or employ multi-wavelength correction could substantially mitigate the melanin interaction. The data presented here provide the quantitative targets for such improvements — and suggest that the engineering problem is more tractable than a simple “melanin blocks light” framing would imply.

### Causal framework

The hypothesized causal structure linking these observations is as follows (see Supplement S7 for directed acyclic graph): critical illness reduces perfusion; low perfusion in the presence of melanin degrades the PPG signal (AC/DC ratio); degraded signal quality produces SpO_2_ overestimation (bias); overestimation produces hidden hypoxemia events (false reassurance); false reassurance may delay clinical escalation; delayed escalation may contribute to mortality. Importantly, illness severity is an upstream common cause of both low perfusion (which drives the signal failure) and mortality (independent of device error). Our severity adjustment and lactate stratification address this confounding path, but we acknowledge that adjustment for downstream mediators (e.g., creatinine and lactate, which are themselves consequences of hypoxia) does not fully resolve the causal structure. This is a fundamental limitation of observational severity adjustment that cannot be overcome without an interventional design.

### 4.4 Clinical Implications: Failure to Rescue

The convergence of the severity adjustment, temporal analysis, and mechanistic data reframes the clinical interpretation of hidden hypoxemia from “statistical noise” to a plausible **failure-to-rescue mechanism.**

The conventional concern is that pulse oximetry bias is merely an artifact of poor circulation at the end of life — that patients were already dying, and the device’s inaccuracy is a symptom, not a cause. Our temporal analyses refute this interpretation. Hidden hypoxemia occurs at a constant rate throughout the ICU stay (3.4–3.8%), not in a pre-terminal surge. The median first hidden event occurs at **15 hours** after ICU admission — during the initial stabilization phase, when ventilator settings are being titrated, oxygen targets are being set, and escalation decisions are being made. Death, when it comes, occurs on average **6.3 days later.** This 150-hour window represents a prolonged interval during which false reassurance from the pulse oximeter could plausibly influence clinical decisions.

In multivariable analysis adjusting for age, sex, race, and four laboratory severity markers, hidden hypoxemia remained independently associated with 86% higher odds of death (OR 1.86, 95% CI 1.69–2.04). The mortality differential (52–103%) substantially exceeds the severity differential (17–41%). And in the landmark analysis — restricting classification to events occurring in the first 48 hours only — the association persisted (Caucasian: 1.88×, African American: 1.55×), confirming that early monitoring failure predicts late mortality.

### The 135-Hour Window for Intervention

Because the initial signal degradation occurs during the median first 15 hours of care — the critical stabilization phase of ICU admission — clinicians are provided inaccurate targets for oxygen titration, ventilator management, and escalation decisions from the outset. The 6.3-day (151-hour) lag between the first hidden event and death defines a concrete window during which automated detection and clinical correction could alter the trajectory. If the pulse oximeter displays 94% while the patient’s true saturation is 85%, the clinical team may not escalate oxygen delivery, may set ventilator targets too low, and may delay recognition of deterioration. Over the ensuing days, this compounding under-treatment may contribute to organ dysfunction and death.

We cannot prove this causal chain in an observational study. But four lines of evidence collectively support it:

1. **Temporal sequence:** The device fails at hour 15; the patient dies at day 6. The error precedes the outcome by 135 hours.
2. **Landmark control:** Restricting to first-48-hour events (uniform exposure window) preserves the ∼1.9× mortality association.
3. **Perfusion independence:** In patients with normal lactate (<2.0 mmol/L) — no shock, no circulatory collapse — hidden hypoxemia remains independently associated with 87% higher odds of death (adjusted OR 1.87, 95% CI 1.61–2.16).
4. **Combined control:** When both the landmark and lactate restrictions are applied simultaneously, mortality remains elevated (16.5% vs. 11.1%).

The 135-hour gap between first failure and death is not an academic abstraction — it is a clinically actionable interval during which the monitoring error could, in principle, be identified and corrected.

African American patients experience 33% more hidden hypoxemia events per ICU stay than Caucasian patients. This represents a measurable, device-mediated disparity in monitoring quality — one that occurs disproportionately during the critical initial hours of care.

### Interpretive scope

These findings should be interpreted as evidence of differential risk stratification and delayed recognition rather than proof that correcting pulse oximetry bias would directly reduce mortality. As an observational analysis, this study cannot establish causal mediation between hidden hypoxemia and death. The observed association instead identifies a prolonged interval during which biased monitoring may plausibly contribute to delayed escalation of care. Whether interventions targeting this interval — such as perfusion-aware algorithms, automated bias flagging, or dual-verification protocols — would reduce mortality is a question that requires prospective evaluation.

### 4.5 Regulatory Relevance and Practical Implications

The demographic variance documented here identifies a **structural limitation in current pulse oximetry technology** rather than a failure of any single manufacturer. The perfusion × melanin interaction is a property of the physics of light absorption through tissue; it affects all devices that rely on dual-wavelength photoplethysmography without perfusion-aware correction. Regulatory standards should accordingly consider complementing warm-subject, healthy-volunteer validation with **multicenter, low-perfusion, population-scale evaluation** that captures the conditions under which the technology fails.

The FDA’s Quality Management System Regulation (QMSR; 21 CFR 820), effective February 2, 2026, requires risk-based quality management and evidence of safety across intended populations^4^. The EU Medical Device Regulation requires clinical evidence from diverse populations. Our data show that multiple subgroups exceed commonly cited performance benchmarks under real-world ICU conditions.

We propose the following framework for translating these findings into practice:

1. **Post-market monitoring:** Population-scale demographic surveillance of pulse oximetry accuracy, using the methodology described herein, as a complement to pre-market validation.
2. **Threshold-based triggers:** When subgroup bias in a multicenter analysis exceeds a defined threshold (e.g., mean bias > 3% in any demographic stratum, or hidden hypoxemia rate > 5% in low-perfusion conditions), manufacturers could use such thresholds as triggers for internal investigation and potential remediation under existing risk-management frameworks. The specific thresholds proposed here are illustrative and would require consensus development.
3. **Pre-submission context:** Manufacturers seeking 510(k) clearance or PMA approval should contextualize their device’s subgroup performance against national baselines such as those presented here.

For example: if a manufacturer’s ongoing surveillance reveals that their device produces a mean bias of +3.5% in African American patients under low-perfusion conditions, this would exceed both the regulatory performance target and the national baseline (+2.76%), warranting investigation and potentially corrective action under QMSR.

These findings are intended to inform research prioritization and the development of surveillance methodologies. They do not imply noncompliance of any specific device or manufacturer, nor do they constitute a determination of regulatory status.

### 4.6 Limitations

This study has several limitations.

First, all analyses are retrospective and observational. The mortality association with hidden hypoxemia is correlational; residual confounding cannot be excluded despite multivariable adjustment. In particular, hidden hypoxemia events could in principle reflect pre-terminal circulatory failure rather than a cause of harm. However, our temporal analyses (Section 3.3.6) argue against this: hidden event rates were constant throughout the ICU stay (not concentrated pre-terminally), the landmark analysis restricted to first-48-hour events preserved the mortality association, the median first hidden event occurred at 15 hours after admission with death following 6.3 days later, and 63% of hidden events occurred more than 48 hours before death. Nonetheless, a randomized trial of monitoring strategies would be required to definitively establish causality.

Additionally, code status (DNR/DNI) was not available in the analyzed datasets. Racial disparities in code status are well-documented in U.S. ICUs, and differential rates of DNR/DNI orders could confound the mortality association. However, we note that (a) the mortality association was consistent across all racial groups, not limited to groups with known higher DNR rates; (b) the adjusted model controlled for multiple severity markers that correlate with end-of-life decisions; and (c) the 86% adjusted odds ratio is large relative to published DNR-associated mortality differences, making it unlikely that code status alone accounts for the observed association. Nonetheless, this remains an unmeasured confounder.

Second, device model and manufacturer information is not available in any database. We characterize **population-level performance of the ecosystem of pulse oximeters deployed in U.S. ICUs**, not device-specific performance. This is a strength (national generalizability) and a limitation (no manufacturer-specific findings).

Third, pairing criteria introduce imprecision. Clinical changes within the 30–60 minute pairing window could affect observed bias. Sensitivity analyses with narrower windows (15 minutes) were consistent with the primary analysis (Supplement S3).

Fourth, the waveform signal quality analysis is based on 125 of 1,545 available subjects. While this is sufficient to demonstrate the absence of a large, fixed racial difference in AC/DC ratio, segment-level analysis correlating AC/DC with concurrent perfusion and SpO_2_ bias — which would directly test the conditional mechanism — requires the full extraction (in progress).

Fifth, the mortality analysis included only ICU stays with ≥1 paired reading (13.7% of MIMIC-IV stays, 27.5% of eICU stays). Stays with paired data may differ systematically from those without (e.g., patients receiving more frequent arterial blood gases may be sicker). Demographic comparisons between included and excluded stays are provided in Supplement S4.

Sixth, the Open Oximetry Repository melanin index is spectrophotometry-derived and may not correspond precisely to clinical or Fitzpatrick classifications. However, as a continuous, objective measure of melanin concentration, it is arguably more informative for optical analysis than categorical race.

Seventh, we did not compute formal Arms (root mean square accuracy) values, which are the standard regulatory metric. Our mean bias estimates under operational conditions are not directly comparable to manufacturer-reported Arms under controlled protocols. We report mean bias as more interpretable for the population-level surveillance framing of this study.

## 5. Conclusion

In this multicenter retrospective analysis of 52 million paired readings across 209 U.S. hospitals, national pulse oximetry variance in African American patients (+2.76%) exceeded prior academic estimates by 62%. Hidden hypoxemia was associated with approximately doubled ICU mortality across all racial and ethnic groups, with an adjusted odds ratio of 1.86 (95% CI 1.69–2.04) after controlling for illness severity. This was not a pre-terminal artifact: hidden events occurred at a constant rate throughout the ICU stay, the first event typically occurred within 15 hours of admission, death followed on average 6.3 days later, and 63% of events occurred more than 48 hours before death. The association persisted in a landmark analysis restricted to first-48-hour events, in patients with normal lactate (adjusted OR 1.87, 95% CI 1.61–2.16), and when both controls were applied simultaneously.

The mechanism is physical and conditional: melanin absorption compounds with low perfusion and hypoxia to produce substantial overestimation (+12.8% in the critical scenario), and the signal deficit emerges dynamically under physiological stress rather than as a fixed baseline property. This implies that the bias is addressable through perfusion-aware algorithms.

We propose the methodology and baselines described herein — the Demographic Oximetry Reference (DOR) — as a framework for ongoing multicenter evaluation of demographic subgroup performance in pulse oximetry. The data, code, and queries are provided in supplementary materials for full reproducibility.

## Supporting information

Gehring_2026_PulseOx_Supplements.zip

## Data Availability

All data used in this study are de-identified and publicly available via PhysioNet (https://physionet.org/). This includes the MIMIC-IV (v3.1), eICU Collaborative Research Database, and the Open Oximetry Repository. Access is provided to credentialed researchers who have completed the required CITI training and signed a Data Use Agreement.

## Data Availability

All source databases are available through PhysioNet (physionet.org) with appropriate credentialing. The complete analytical pipeline — from raw SQL queries to Python-based signal quality algorithms — is open-source and will be hosted in a public repository [URL to be added upon submission]. Every query that produced every number in this manuscript is provided in Supplement S1, and the intermediate BigQuery tables are available for inspection (project: candor-health-486219, dataset: algorithm_benchmarks). Independent replication and audit of these analyses is encouraged and is enabled through the credentialed access protocols of PhysioNet. No data were filtered, excluded, or transformed beyond the explicit criteria described in Section 2.2.

## Funding and Compute Resources

This research was conducted independently without traditional grant funding. Cloud computing infrastructure was provided through the Google Cloud Platform (GCP) Startup program, which supported all BigQuery analyses and Vertex AI workloads. Additional compute resources were provided by NVIDIA through the Inception program and by Lambda Labs. Amazon Web Services (AWS) provided supplementary cloud credits for data transfer operations. The funders had no role in study design, data analysis, interpretation, or manuscript preparation.

## Acknowledgements

We gratefully acknowledge PhysioNet and the research teams who created and maintain the clinical databases that made this work possible: the MIMIC team (Alistair Johnson, Tom Pollard, Lucas Bulgarelli, and colleagues at the MIT Laboratory for Computational Physiology and Beth Israel Deaconess Medical Center); the eICU Collaborative Research Database team; the Open Oximetry Repository contributors; and the MIMIC-III Waveform Database authors. The commitment of these groups to open, credentialed access to clinical data is essential to independent research on device safety and health equity. We also acknowledge the patients whose deidentified data underlie these analyses.

## Conflicts of Interest

MG is the founder of Candor Systems, which provides demographic bias evaluation services for medical device manufacturers. The analysis presented here was developed independently using publicly available clinical databases under standard PhysioNet credentialed access. No proprietary data, algorithms, or manufacturer relationships influenced any aspect of the study design, analysis, or interpretation. The complete analytical code is provided for independent verification.

## Author Contributions

MG conceived the study, obtained data access, designed and executed all analyses, and wrote the manuscript. External clinical and statistical experts reviewed the analytic framework and interpretation prior to submission.

## Supplementary Materials

*Available online:*

- **S1:** Complete SQL queries for all analyses (full reproducibility)
- **S2:** Python code for waveform feature extraction (golden_subset_pipeline.py)
- **S3:** Sensitivity analyses (pairing windows, thresholds, patient-level aggregation)
- **S4:** Missingness analysis — demographic comparisons of included vs. excluded stays
- **S5:** Full multivariable logistic regression model for mortality (coefficients, ORs, 95% CIs)
- **S6:** Dataset access instructions and reproducibility guide
- **S7:** Directed acyclic graph (DAG) — hypothesized causal structure linking illness severity, perfusion, melanin, signal quality, bias, hidden hypoxemia, and mortality

## Notes

### Competing Interest Statement

Matthew Gehring is the founder of Candor Systems, which provides demographic bias evaluation services for medical device manufacturers. The analysis was developed independently using publicly available clinical databases under standard PhysioNet credentialed access. No proprietary data, algorithms, or manufacturer relationships influenced any aspect of the study design, analysis, or interpretation.

### Funding Statement

This study did not receive any funding.

### Author Declarations

This study utilized de-identified, publicly available electronic health record data from the PhysioNet platform (MIMIC-IV, eICU-CRD, and the Open Oximetry Repository). Access was granted following completion of the CITI Program 'Data or Specimens Only Research' training. According to the U.S. Department of Health and Human Services (HHS) regulations (45 CFR 46), the use of these de-identified datasets does not constitute human subjects research and is exempt from Institutional Review Board (IRB) review.

