## Supplementary material for "Perfusion-Dependent Melanin Bias in Pulse Oximetry and ICU Mortality Across 209 U.S. Hospitals: A Multicenter Retrospective Analysis of 52 Million Readings": Gehring_2026_PulseOx_Supplements.zip: LICENSE

Creative Commons Attribution 4.0 International (CC BY 4.0)

Copyright (c) 2026 Matthew Gehring / Candor Systems

You are free to:

  Share — copy and redistribute the material in any medium or format
  Adapt — remix, transform, and build upon the material for any purpose,
          even commercially

Under the following terms:

  Attribution — You must give appropriate credit, provide a link to the
  license, and indicate if changes were made. You may do so in any
  reasonable manner, but not in any way that suggests the licensor
  endorses you or your use.

Full license text: https://creativecommons.org/licenses/by/4.0/legalcode
