## Supplementary material for "Perfusion-Dependent Melanin Bias in Pulse Oximetry and ICU Mortality Across 209 U.S. Hospitals: A Multicenter Retrospective Analysis of 52 Million Readings": Gehring_2026_PulseOx_Supplements.zip: README.md

# Supplementary Materials

**Perfusion-Dependent Melanin Bias in Pulse Oximetry and ICU Mortality Across 209 U.S. Hospitals: A Multicenter Retrospective Analysis of 52 Million Readings**

Matthew Gehring, Candor Systems

Preprint: Submitted to medRxiv (February 2026).

---

## Overview

This repository contains the complete supplementary materials for the above manuscript. Every number reported in the paper can be independently reproduced from these files using publicly available PhysioNet databases and Google BigQuery.

---

## File index

| File | Supplement | Description |
|------|-----------|-------------|
| `Supplement_S1_SQL_Queries.sql` | S1 | All SQL queries (BigQuery Standard SQL) organized by manuscript section. Parts A–G cover bias estimation, regression input, sensitivity analyses, missingness, temporal/landmark analyses, lactate stratification, and Golden Subset waveform clinical linkage. |
| `Supplement_S2_Waveform_Extraction.py` | S2 | Python pipeline for MIMIC-III waveform inventory and PPG signal-quality feature extraction (AC/DC ratio, SNR, entropy). Designed for Vertex AI / any GCS-connected environment. |
| `Supplement_S3_Sensitivity_Analyses.md` | S3 | Formatted results tables: pairing window (15 min), alternate hidden-hypoxemia thresholds (92/90), patient-level aggregated bias, and minimum-3-readings restriction. |
| `Supplement_S4_Missingness_Analysis.md` | S4 | Demographic comparison (age, sex, ethnicity, mortality) of ICU stays included vs. excluded from the mortality analysis (eICU-CRD). |
| `Supplement_S5_Full_Regression_Model.md` | S5 | Full logistic regression output: coefficients, standard errors, p-values, odds ratios, and 95% CIs for both the full-cohort model (OR 1.86) and the normal-lactate subgroup (OR 1.87), with cluster-robust SEs by hospital. |
| `Supplement_S6_Reproducibility_Guide.md` | S6 | Step-by-step instructions: obtain PhysioNet credentialed access, configure BigQuery, run S1 queries, execute S2 pipeline, and reproduce the logistic regression in Python (statsmodels). |
| `Supplement_S7_Causal_DAG.md` | S7 | Text description and minimal ASCII diagram of the directed acyclic graph: nodes (illness severity, perfusion, melanin, PPG signal quality, bias, hidden hypoxemia, delayed escalation, mortality), directed edges, and implications for confounder/mediator adjustment. |
| `LICENSE` | — | CC BY 4.0 International. |

---

## Data availability

All underlying clinical data are hosted on PhysioNet and require credentialed access (free):

- **MIMIC-IV v3.1** — `physionet-data.mimiciv_3_1_hosp` / `mimiciv_3_1_icu`
- **eICU-CRD** — `physionet-data.eicu_crd`
- **Open Oximetry Repository** — PhysioNet (openoximetry, v1.0)
- **MIMIC-III Clinical** — `physionet-data.mimiciii_clinical`
- **MIMIC-III Waveform** — PhysioNet (mimic3wdb)

See `Supplement_S6_Reproducibility_Guide.md` for detailed access instructions.

---

## How to cite

If you use these materials, please cite both the manuscript and this deposit:

> Gehring M. Supplementary materials for "Perfusion-Dependent Melanin Bias in Pulse Oximetry and ICU Mortality Across 209 U.S. Hospitals." Zenodo. 2026. DOI: 10.5281/zenodo.18530195

> Gehring M. Perfusion-Dependent Melanin Bias in Pulse Oximetry and ICU Mortality Across 209 U.S. Hospitals: A Multicenter Retrospective Analysis of 52 Million Readings. Submitted to medRxiv. 2026.

---

## License

This work is licensed under a [Creative Commons Attribution 4.0 International License](https://creativecommons.org/licenses/by/4.0/).

---

## Contact

Matthew Gehring —
