## Supplementary material for "Perfusion-Dependent Melanin Bias in Pulse Oximetry and ICU Mortality Across 209 U.S. Hospitals: A Multicenter Retrospective Analysis of 52 Million Readings": Gehring_2026_PulseOx_Supplements.zip: Supplement_S1_SQL_Queries.sql

```
-- =============================================================================
-- SUPPLEMENT S1 — Complete SQL Queries for Reproducibility
-- Gehring 2026: Perfusion-Dependent Melanin Bias in Pulse Oximetry and ICU
--              Mortality Across 209 U.S. Hospitals (Multicenter Retrospective)
-- =============================================================================
-- Each section maps to manuscript Tables/Figures. Run in BigQuery (Standard SQL).
-- Datasets: physionet-data (MIMIC-IV: mimiciv_3_1_hosp/icu; eICU: eicu_crd;
--           MIMIC-III: mimiciii_clinical); candor-health-486219.algorithm_benchmarks
-- Prerequisites: PhysioNet credentialed access; table mimic3_waveform_subjects
--                for Golden Subset sections (create via pipeline or BQ cohort).
-- =============================================================================


-- =============================================================================
-- PART A: BIAS AND DEMOGRAPHICS (Manuscript Table 2 — eICU patient-level bias)
-- =============================================================================
-- Query A1: Bias estimates with SD and 95% CI by ethnicity (eICU).
-- Output: Table 2 panel (patient-level), Section 3.2.

WITH patient_bias AS (
    SELECT
        pt.ethnicity,
        v.patientunitstayid,
        AVG(SAFE_CAST(v.sao2 AS FLOAT64) - SAFE_CAST(l.labresult AS FLOAT64)) AS mean_bias_per_stay
    FROM `physionet-data.eicu_crd.vitalperiodic` v
    JOIN `physionet-data.eicu_crd.patient` pt ON v.patientunitstayid = pt.patientunitstayid
    JOIN `physionet-data.eicu_crd.lab` l ON l.patientunitstayid = v.patientunitstayid
        AND l.labname = 'O2 Sat (%)'
        AND ABS(l.labresultoffset - v.observationoffset) <= 60
    WHERE v.sao2 IS NOT NULL
        AND SAFE_CAST(v.sao2 AS FLOAT64) BETWEEN 50 AND 100
        AND SAFE_CAST(l.labresult AS FLOAT64) BETWEEN 50 AND 100
    GROUP BY 1, 2
)
SELECT
    ethnicity,
    COUNT(*) AS n_stays,
    ROUND(AVG(mean_bias_per_stay), 3) AS mean_bias,
    ROUND(STDDEV(mean_bias_per_stay), 3) AS sd_bias,
    ROUND(AVG(mean_bias_per_stay) - 1.96 * STDDEV(mean_bias_per_stay) / SQRT(COUNT(*)), 3) AS ci_lower,
    ROUND(AVG(mean_bias_per_stay) + 1.96 * STDDEV(mean_bias_per_stay) / SQRT(COUNT(*)), 3) AS ci_upper
FROM patient_bias
GROUP BY 1
ORDER BY 1;


-- =============================================================================
-- PART B: REGRESSION INPUT (Manuscript Table 6c, Supplement S5)
-- =============================================================================
-- Query B1: Creates per-stay table with outcome and covariates for logistic
-- regression (full cohort). Export to CSV and run in Python/R with cluster-robust
-- SEs by hospital. See S5 for model specification and results.

CREATE OR REPLACE TABLE `candor-health-486219.algorithm_benchmarks.eicu_regression_input` AS
WITH paired AS (
    SELECT
        v.patientunitstayid,
        pt.ethnicity,
        pt.gender,
        pt.age,
        pt.hospitaldischargestatus,
        SAFE_CAST(v.sao2 AS FLOAT64) AS spo2_monitor,
        SAFE_CAST(l.labresult AS FLOAT64) AS sao2_lab,
        CASE WHEN SAFE_CAST(v.sao2 AS FLOAT64) >= 94
              AND SAFE_CAST(l.labresult AS FLOAT64) < 88 THEN 1 ELSE 0 END AS hidden_severe
    FROM `physionet-data.eicu_crd.vitalperiodic` v
    JOIN `physionet-data.eicu_crd.patient` pt ON v.patientunitstayid = pt.patientunitstayid
    JOIN `physionet-data.eicu_crd.lab` l ON l.patientunitstayid = v.patientunitstayid
        AND l.labname = 'O2 Sat (%)'
        AND ABS(l.labresultoffset - v.observationoffset) <= 60
    WHERE v.sao2 IS NOT NULL
        AND SAFE_CAST(v.sao2 AS FLOAT64) BETWEEN 50 AND 100
        AND SAFE_CAST(l.labresult AS FLOAT64) BETWEEN 50 AND 100
),
per_stay AS (
    SELECT
        patientunitstayid,
        ANY_VALUE(ethnicity) AS ethnicity,
        ANY_VALUE(gender) AS gender,
        ANY_VALUE(age) AS age,
        ANY_VALUE(hospitaldischargestatus) AS discharge_status,
        COUNT(*) AS n_paired,
        SUM(hidden_severe) AS hidden_count,
        CASE WHEN SUM(hidden_severe) > 0 THEN 1 ELSE 0 END AS hidden_flag
    FROM paired
    GROUP BY 1
),
severity AS (
    SELECT
        patientunitstayid,
        MAX(CASE WHEN labname = 'creatinine' THEN SAFE_CAST(labresult AS FLOAT64) END) AS max_creatinine,
        MAX(CASE WHEN labname = 'total bilirubin' THEN SAFE_CAST(labresult AS FLOAT64) END) AS max_bilirubin,
        MIN(CASE WHEN labname = 'platelets x 1000' THEN SAFE_CAST(labresult AS FLOAT64) END) AS min_platelets,
        MAX(CASE WHEN labname = 'lactate' THEN SAFE_CAST(labresult AS FLOAT64) END) AS max_lactate
    FROM `physionet-data.eicu_crd.lab`
    GROUP BY 1
),
hospital AS (
    SELECT patientunitstayid, hospitalid
    FROM `physionet-data.eicu_crd.patient`
)
SELECT
    ps.patientunitstayid,
    h.hospitalid,
    ps.ethnicity,
    ps.gender,
    SAFE_CAST(ps.age AS FLOAT64) AS age,
    ps.hidden_flag,
    ps.hidden_count,
    ps.n_paired,
    CASE WHEN ps.discharge_status = 'Expired' THEN 1 ELSE 0 END AS died,
    s.max_creatinine,
    s.max_bilirubin,
    s.min_platelets,
    s.max_lactate
FROM per_stay ps
LEFT JOIN severity s ON ps.patientunitstayid = s.patientunitstayid
LEFT JOIN hospital h ON ps.patientunitstayid = h.patientunitstayid;


-- =============================================================================
-- PART C: SENSITIVITY ANALYSES (Supplement S3)
-- =============================================================================
-- Query C1: Pairing window 15 min (MIMIC-IV) — mortality by hidden status.
-- Query C2: Alternate threshold SpO2≥92, SaO2<90 (eICU).
-- Query C3: Patient-level aggregated bias (eICU) — Table 2 / S3.
-- Query C4: Minimum 3 paired readings per stay (eICU).

-- C1: 15-min pairing (MIMIC-IV)
WITH spo2 AS (
    SELECT SUBJECT_ID, HADM_ID, CHARTTIME, VALUENUM AS spo2
    FROM `physionet-data.mimiciv_3_1_icu.chartevents`
    WHERE ITEMID = 220277 AND VALUENUM BETWEEN 50 AND 100
),
sao2 AS (
    SELECT SUBJECT_ID, HADM_ID, CHARTTIME, VALUENUM AS sao2
    FROM `physionet-data.mimiciv_3_1_icu.chartevents`
    WHERE ITEMID = 220227 AND VALUENUM BETWEEN 50 AND 100
),
paired_15 AS (
    SELECT
        s.SUBJECT_ID, s.HADM_ID,
        s.spo2, a.sao2,
        s.spo2 - a.sao2 AS bias,
        CASE WHEN s.spo2 >= 94 AND a.sao2 < 88 THEN 1 ELSE 0 END AS hidden_severe,
        ROW_NUMBER() OVER (PARTITION BY s.SUBJECT_ID, s.HADM_ID, s.CHARTTIME
            ORDER BY ABS(DATETIME_DIFF(s.CHARTTIME, a.CHARTTIME, MINUTE))) AS rn
    FROM spo2 s
    JOIN sao2 a ON s.SUBJECT_ID = a.SUBJECT_ID AND s.HADM_ID = a.HADM_ID
        AND ABS(DATETIME_DIFF(s.CHARTTIME, a.CHARTTIME, MINUTE)) <= 15
),
per_stay AS (
    SELECT
        p.SUBJECT_ID, p.HADM_ID,
        CASE WHEN SUM(p.hidden_severe) > 0 THEN 'Hidden' ELSE 'No hidden' END AS hidden_group
    FROM paired_15 p
    WHERE p.rn = 1
    GROUP BY 1, 2
)
SELECT
    adm.race,
    ps.hidden_group,
    COUNT(*) AS n_stays,
    SUM(adm.hospital_expire_flag) AS deaths,
    ROUND(100.0 * SUM(adm.hospital_expire_flag) / COUNT(*), 1) AS mortality_pct
FROM per_stay ps
JOIN `physionet-data.mimiciv_3_1_hosp.admissions` adm
    ON ps.SUBJECT_ID = adm.subject_id AND ps.HADM_ID = adm.hadm_id
WHERE adm.race IN ('WHITE', 'BLACK/AFRICAN AMERICAN')
GROUP BY 1, 2
ORDER BY 1, 2;

-- C2: Alternate threshold 92/90 (eICU)
WITH paired AS (
    SELECT
        v.patientunitstayid,
        pt.ethnicity,
        pt.hospitaldischargestatus,
        SAFE_CAST(v.sao2 AS FLOAT64) AS spo2_monitor,
        SAFE_CAST(l.labresult AS FLOAT64) AS sao2_lab,
        CASE WHEN SAFE_CAST(v.sao2 AS FLOAT64) >= 92
              AND SAFE_CAST(l.labresult AS FLOAT64) < 90 THEN 1 ELSE 0 END AS hidden_alt
    FROM `physionet-data.eicu_crd.vitalperiodic` v
    JOIN `physionet-data.eicu_crd.patient` pt ON v.patientunitstayid = pt.patientunitstayid
    JOIN `physionet-data.eicu_crd.lab` l ON l.patientunitstayid = v.patientunitstayid
        AND l.labname = 'O2 Sat (%)'
        AND ABS(l.labresultoffset - v.observationoffset) <= 60
    WHERE v.sao2 IS NOT NULL
        AND SAFE_CAST(v.sao2 AS FLOAT64) BETWEEN 50 AND 100
        AND SAFE_CAST(l.labresult AS FLOAT64) BETWEEN 50 AND 100
),
per_stay AS (
    SELECT
        patientunitstayid, ethnicity, hospitaldischargestatus,
        CASE WHEN SUM(hidden_alt) > 0 THEN 'Hidden (92/90)' ELSE 'No hidden' END AS hidden_group
    FROM paired
    GROUP BY 1, 2, 3
)
SELECT
    ethnicity,
    hidden_group,
    COUNT(*) AS n_stays,
    ROUND(100.0 * COUNTIF(hospitaldischargestatus = 'Expired') / COUNT(*), 1) AS mortality_pct
FROM per_stay
GROUP BY 1, 2
ORDER BY 1, 2;

-- C3: Patient-level aggregated bias (eICU)
WITH patient_bias AS (
    SELECT
        pt.ethnicity,
        v.patientunitstayid,
        AVG(SAFE_CAST(v.sao2 AS FLOAT64) - SAFE_CAST(l.labresult AS FLOAT64)) AS mean_bias
    FROM `physionet-data.eicu_crd.vitalperiodic` v
    JOIN `physionet-data.eicu_crd.patient` pt ON v.patientunitstayid = pt.patientunitstayid
    JOIN `physionet-data.eicu_crd.lab` l ON l.patientunitstayid = v.patientunitstayid
        AND l.labname = 'O2 Sat (%)'
        AND ABS(l.labresultoffset - v.observationoffset) <= 60
    WHERE v.sao2 IS NOT NULL
        AND SAFE_CAST(v.sao2 AS FLOAT64) BETWEEN 50 AND 100
        AND SAFE_CAST(l.labresult AS FLOAT64) BETWEEN 50 AND 100
    GROUP BY 1, 2
)
SELECT
    ethnicity,
    COUNT(*) AS n_stays,
    ROUND(AVG(mean_bias), 3) AS mean_bias_patient_level,
    ROUND(STDDEV(mean_bias), 3) AS sd,
    ROUND(APPROX_QUANTILES(mean_bias, 4)[OFFSET(2)], 3) AS median_bias
FROM patient_bias
GROUP BY 1
ORDER BY 1;

-- C4: Minimum 3 paired readings per stay (eICU)
WITH paired AS (
    SELECT
        v.patientunitstayid,
        pt.ethnicity,
        pt.hospitaldischargestatus,
        CASE WHEN SAFE_CAST(v.sao2 AS FLOAT64) >= 94
              AND SAFE_CAST(l.labresult AS FLOAT64) < 88 THEN 1 ELSE 0 END AS hidden_severe
    FROM `physionet-data.eicu_crd.vitalperiodic` v
    JOIN `physionet-data.eicu_crd.patient` pt ON v.patientunitstayid = pt.patientunitstayid
    JOIN `physionet-data.eicu_crd.lab` l ON l.patientunitstayid = v.patientunitstayid
        AND l.labname = 'O2 Sat (%)'
        AND ABS(l.labresultoffset - v.observationoffset) <= 60
    WHERE v.sao2 IS NOT NULL
        AND SAFE_CAST(v.sao2 AS FLOAT64) BETWEEN 50 AND 100
        AND SAFE_CAST(l.labresult AS FLOAT64) BETWEEN 50 AND 100
),
per_stay AS (
    SELECT
        patientunitstayid, ethnicity, hospitaldischargestatus,
        COUNT(*) AS n_paired,
        SUM(hidden_severe) AS hidden_count,
        CASE WHEN SUM(hidden_severe) > 0 THEN 'Hidden' ELSE 'No hidden' END AS hidden_group
    FROM paired
    GROUP BY 1, 2, 3
)
SELECT
    ethnicity,
    hidden_group,
    COUNT(*) AS n_stays,
    ROUND(100.0 * COUNTIF(hospitaldischargestatus = 'Expired') / COUNT(*), 1) AS mortality_pct
FROM per_stay
WHERE n_paired >= 3
GROUP BY 1, 2
ORDER BY 1, 2;


-- =============================================================================
-- PART D: MISSINGNESS (Supplement S4 — Manuscript Section 2.11)
-- =============================================================================
-- Query D1: Demographics of included vs excluded stays (eICU).

WITH has_paired AS (
    SELECT DISTINCT v.patientunitstayid
    FROM `physionet-data.eicu_crd.vitalperiodic` v
    JOIN `physionet-data.eicu_crd.lab` l ON l.patientunitstayid = v.patientunitstayid
        AND l.labname = 'O2 Sat (%)'
        AND ABS(l.labresultoffset - v.observationoffset) <= 60
    WHERE v.sao2 IS NOT NULL
        AND SAFE_CAST(v.sao2 AS FLOAT64) BETWEEN 50 AND 100
        AND SAFE_CAST(l.labresult AS FLOAT64) BETWEEN 50 AND 100
)
SELECT
    CASE WHEN hp.patientunitstayid IS NOT NULL THEN 'Included' ELSE 'Excluded' END AS group_label,
    pt.ethnicity,
    COUNT(*) AS n_stays,
    ROUND(AVG(SAFE_CAST(pt.age AS FLOAT64)), 1) AS mean_age,
    ROUND(100.0 * COUNTIF(pt.gender = 'Male') / COUNT(*), 1) AS pct_male,
    ROUND(100.0 * COUNTIF(pt.hospitaldischargestatus = 'Expired') / COUNT(*), 1) AS mortality_pct
FROM `physionet-data.eicu_crd.patient` pt
LEFT JOIN has_paired hp ON pt.patientunitstayid = hp.patientunitstayid
GROUP BY 1, 2
ORDER BY 1, 2;


-- =============================================================================
-- PART E: TEMPORAL / LANDMARK (Manuscript Tables 6d–6g, Section 3.3.6)
-- =============================================================================
-- E1: Hidden event rate by time window (Table 6d).
-- E2: Landmark — first-48h hidden hypoxemia → mortality (Table 6e).
-- E3: Timing of first hidden event (Table 6f).
-- E4: Distribution of hidden events by hours before death (Table 6g).

-- E1: Time window
WITH paired AS (
    SELECT v.patientunitstayid,
        pt.ethnicity,
        pt.hospitaldischargestatus,
        v.observationoffset,
        SAFE_CAST(v.sao2 AS FLOAT64) AS spo2_monitor,
        SAFE_CAST(l.labresult AS FLOAT64) AS sao2_lab,
        CASE WHEN SAFE_CAST(v.sao2 AS FLOAT64) >= 94 AND SAFE_CAST(l.labresult AS FLOAT64) < 88 THEN 1 ELSE 0 END AS hidden_severe
    FROM `physionet-data.eicu_crd.vitalperiodic` v
    JOIN `physionet-data.eicu_crd.patient` pt ON v.patientunitstayid = pt.patientunitstayid
    JOIN `physionet-data.eicu_crd.lab` l ON l.patientunitstayid = v.patientunitstayid
        AND l.labname = 'O2 Sat (%)'
        AND ABS(l.labresultoffset - v.observationoffset) <= 60
    WHERE v.sao2 IS NOT NULL
        AND SAFE_CAST(v.sao2 AS FLOAT64) BETWEEN 50 AND 100
        AND SAFE_CAST(l.labresult AS FLOAT64) BETWEEN 50 AND 100
)
SELECT
    CASE
        WHEN observationoffset <= 1440 THEN '1_First 24h'
        WHEN observationoffset <= 2880 THEN '2_24-48h'
        WHEN observationoffset <= 4320 THEN '3_48-72h'
        ELSE '4_After 72h'
    END AS time_window,
    COUNT(*) AS n_paired_readings,
    SUM(hidden_severe) AS n_hidden_severe,
    ROUND(100.0 * SUM(hidden_severe) / COUNT(*), 2) AS pct_hidden_severe
FROM paired
GROUP BY 1
ORDER BY 1;

-- E2: Landmark first-48h
WITH paired AS (
    SELECT v.patientunitstayid,
        pt.ethnicity,
        pt.hospitaldischargestatus,
        v.observationoffset,
        CASE WHEN SAFE_CAST(v.sao2 AS FLOAT64) >= 94 AND SAFE_CAST(l.labresult AS FLOAT64) < 88 THEN 1 ELSE 0 END AS hidden_severe
    FROM `physionet-data.eicu_crd.vitalperiodic` v
    JOIN `physionet-data.eicu_crd.patient` pt ON v.patientunitstayid = pt.patientunitstayid
    JOIN `physionet-data.eicu_crd.lab` l ON l.patientunitstayid = v.patientunitstayid
        AND l.labname = 'O2 Sat (%)'
        AND ABS(l.labresultoffset - v.observationoffset) <= 60
    WHERE v.sao2 IS NOT NULL
        AND SAFE_CAST(v.sao2 AS FLOAT64) BETWEEN 50 AND 100
        AND SAFE_CAST(l.labresult AS FLOAT64) BETWEEN 50 AND 100
        AND v.observationoffset <= 2880
),
per_stay AS (
    SELECT patientunitstayid,
        ANY_VALUE(ethnicity) AS ethnicity,
        ANY_VALUE(hospitaldischargestatus) AS discharge_status,
        SUM(hidden_severe) AS hidden_count,
        CASE WHEN SUM(hidden_severe) > 0 THEN 'Hidden (first 48h)' ELSE 'No hidden (first 48h)' END AS hidden_group
    FROM paired
    GROUP BY 1
)
SELECT ethnicity, hidden_group,
    COUNT(*) AS n_stays,
    ROUND(100.0 * COUNTIF(discharge_status = 'Expired') / COUNT(*), 1) AS mortality_pct
FROM per_stay
GROUP BY 1, 2
ORDER BY 1, 2;

-- E3: First hidden event timing
WITH paired AS (
    SELECT v.patientunitstayid,
        pt.hospitaldischargestatus,
        pt.hospitaldischargeoffset,
        v.observationoffset,
        CASE WHEN SAFE_CAST(v.sao2 AS FLOAT64) >= 94 AND SAFE_CAST(l.labresult AS FLOAT64) < 88 THEN 1 ELSE 0 END AS hidden_severe
    FROM `physionet-data.eicu_crd.vitalperiodic` v
    JOIN `physionet-data.eicu_crd.patient` pt ON v.patientunitstayid = pt.patientunitstayid
    JOIN `physionet-data.eicu_crd.lab` l ON l.patientunitstayid = v.patientunitstayid
        AND l.labname = 'O2 Sat (%)'
        AND ABS(l.labresultoffset - v.observationoffset) <= 60
    WHERE v.sao2 IS NOT NULL
        AND SAFE_CAST(v.sao2 AS FLOAT64) BETWEEN 50 AND 100
        AND SAFE_CAST(l.labresult AS FLOAT64) BETWEEN 50 AND 100
),
first_hidden AS (
    SELECT patientunitstayid,
        ANY_VALUE(hospitaldischargestatus) AS discharge_status,
        ANY_VALUE(hospitaldischargeoffset) AS discharge_offset,
        MIN(CASE WHEN hidden_severe = 1 THEN observationoffset END) AS first_hidden_offset_min
    FROM paired
    GROUP BY 1
    HAVING SUM(hidden_severe) > 0
)
SELECT discharge_status,
    COUNT(*) AS n_stays,
    ROUND(AVG(first_hidden_offset_min / 60.0), 1) AS mean_first_hidden_hours,
    ROUND(APPROX_QUANTILES(first_hidden_offset_min / 60.0, 4)[OFFSET(2)], 1) AS median_first_hidden_hours,
    ROUND(AVG(discharge_offset / 60.0), 1) AS mean_los_hours,
    ROUND(AVG((discharge_offset - first_hidden_offset_min) / 60.0), 1) AS mean_hours_from_hidden_to_discharge
FROM first_hidden
GROUP BY 1
ORDER BY 1;

-- E4: Hours before death
WITH paired AS (
    SELECT v.patientunitstayid,
        pt.hospitaldischargestatus,
        pt.hospitaldischargeoffset,
        v.observationoffset,
        CASE WHEN SAFE_CAST(v.sao2 AS FLOAT64) >= 94 AND SAFE_CAST(l.labresult AS FLOAT64) < 88 THEN 1 ELSE 0 END AS hidden_severe
    FROM `physionet-data.eicu_crd.vitalperiodic` v
    JOIN `physionet-data.eicu_crd.patient` pt ON v.patientunitstayid = pt.patientunitstayid
    JOIN `physionet-data.eicu_crd.lab` l ON l.patientunitstayid = v.patientunitstayid
        AND l.labname = 'O2 Sat (%)'
        AND ABS(l.labresultoffset - v.observationoffset) <= 60
    WHERE v.sao2 IS NOT NULL
        AND SAFE_CAST(v.sao2 AS FLOAT64) BETWEEN 50 AND 100
        AND SAFE_CAST(l.labresult AS FLOAT64) BETWEEN 50 AND 100
        AND pt.hospitaldischargestatus = 'Expired'
),
hidden_events AS (
    SELECT patientunitstayid,
        observationoffset,
        hospitaldischargeoffset,
        (hospitaldischargeoffset - observationoffset) / 60.0 AS hours_before_death
    FROM paired
    WHERE hidden_severe = 1
)
SELECT
    CASE
        WHEN hours_before_death <= 2 THEN '1_Final 2h'
        WHEN hours_before_death <= 6 THEN '2_2-6h before death'
        WHEN hours_before_death <= 24 THEN '3_6-24h before death'
        WHEN hours_before_death <= 48 THEN '4_24-48h before death'
        ELSE '5_>48h before death'
    END AS timing_bucket,
    COUNT(*) AS n_hidden_events,
    ROUND(100.0 * COUNT(*) / SUM(COUNT(*)) OVER (), 1) AS pct_of_all_hidden_events
FROM hidden_events
WHERE hours_before_death >= 0
GROUP BY 1
ORDER BY 1;


-- =============================================================================
-- PART F: LACTATE STRATIFICATION (Manuscript Table 6h, Section 3.3.7)
-- =============================================================================
-- F1: Mortality by lactate group and hidden status (Table 6h).
-- F2: Regression input for normal-lactate subgroup (feeds S5).
-- F3: Combined first-48h + normal lactate.

-- F1: Lactate × hidden
WITH paired AS (
    SELECT v.patientunitstayid, pt.ethnicity, pt.hospitaldischargestatus,
        CASE WHEN SAFE_CAST(v.sao2 AS FLOAT64) >= 94 AND SAFE_CAST(l.labresult AS FLOAT64) < 88 THEN 1 ELSE 0 END AS hidden_severe
    FROM `physionet-data.eicu_crd.vitalperiodic` v
    JOIN `physionet-data.eicu_crd.patient` pt ON v.patientunitstayid = pt.patientunitstayid
    JOIN `physionet-data.eicu_crd.lab` l ON l.patientunitstayid = v.patientunitstayid
        AND l.labname = 'O2 Sat (%)'
        AND ABS(l.labresultoffset - v.observationoffset) <= 60
    WHERE v.sao2 IS NOT NULL
        AND SAFE_CAST(v.sao2 AS FLOAT64) BETWEEN 50 AND 100
        AND SAFE_CAST(l.labresult AS FLOAT64) BETWEEN 50 AND 100
),
per_stay AS (
    SELECT patientunitstayid,
        ANY_VALUE(ethnicity) AS ethnicity,
        ANY_VALUE(hospitaldischargestatus) AS discharge_status,
        CASE WHEN SUM(hidden_severe) > 0 THEN 1 ELSE 0 END AS hidden_flag
    FROM paired
    GROUP BY 1
),
lactate AS (
    SELECT patientunitstayid,
        MAX(SAFE_CAST(labresult AS FLOAT64)) AS max_lactate
    FROM `physionet-data.eicu_crd.lab`
    WHERE labname = 'lactate' AND SAFE_CAST(labresult AS FLOAT64) BETWEEN 0.1 AND 50
    GROUP BY 1
)
SELECT
    CASE
        WHEN la.max_lactate < 2.0 THEN '1_Normal lactate (<2.0)'
        WHEN la.max_lactate < 4.0 THEN '2_Elevated (2.0-4.0)'
        WHEN la.max_lactate >= 4.0 THEN '3_Severe (>=4.0)'
        ELSE '4_Missing'
    END AS lactate_group,
    CASE WHEN ps.hidden_flag = 1 THEN 'Hidden' ELSE 'No hidden' END AS hidden_group,
    COUNT(*) AS n_stays,
    ROUND(100.0 * COUNTIF(ps.discharge_status = 'Expired') / COUNT(*), 1) AS mortality_pct
FROM per_stay ps
LEFT JOIN lactate la ON ps.patientunitstayid = la.patientunitstayid
GROUP BY 1, 2
ORDER BY 1, 2;

-- F2: Normal-lactate regression table
CREATE OR REPLACE TABLE `candor-health-486219.algorithm_benchmarks.eicu_normal_lactate_regression` AS
WITH paired AS (
    SELECT v.patientunitstayid, pt.ethnicity, pt.gender, pt.age, pt.hospitaldischargestatus, pt.hospitalid,
        CASE WHEN SAFE_CAST(v.sao2 AS FLOAT64) >= 94 AND SAFE_CAST(l.labresult AS FLOAT64) < 88 THEN 1 ELSE 0 END AS hidden_severe
    FROM `physionet-data.eicu_crd.vitalperiodic` v
    JOIN `physionet-data.eicu_crd.patient` pt ON v.patientunitstayid = pt.patientunitstayid
    JOIN `physionet-data.eicu_crd.lab` l ON l.patientunitstayid = v.patientunitstayid
        AND l.labname = 'O2 Sat (%)'
        AND ABS(l.labresultoffset - v.observationoffset) <= 60
    WHERE v.sao2 IS NOT NULL
        AND SAFE_CAST(v.sao2 AS FLOAT64) BETWEEN 50 AND 100
        AND SAFE_CAST(l.labresult AS FLOAT64) BETWEEN 50 AND 100
),
per_stay AS (
    SELECT patientunitstayid,
        ANY_VALUE(ethnicity) AS ethnicity,
        ANY_VALUE(gender) AS gender,
        ANY_VALUE(age) AS age,
        ANY_VALUE(hospitaldischargestatus) AS discharge_status,
        ANY_VALUE(hospitalid) AS hospitalid,
        CASE WHEN SUM(hidden_severe) > 0 THEN 1 ELSE 0 END AS hidden_flag
    FROM paired
    GROUP BY 1
),
severity AS (
    SELECT patientunitstayid,
        MAX(CASE WHEN labname = 'creatinine' THEN SAFE_CAST(labresult AS FLOAT64) END) AS max_creatinine,
        MAX(CASE WHEN labname = 'total bilirubin' THEN SAFE_CAST(labresult AS FLOAT64) END) AS max_bilirubin,
        MIN(CASE WHEN labname = 'platelets x 1000' THEN SAFE_CAST(labresult AS FLOAT64) END) AS min_platelets,
        MAX(CASE WHEN labname = 'lactate' THEN SAFE_CAST(labresult AS FLOAT64) END) AS max_lactate
    FROM `physionet-data.eicu_crd.lab`
    GROUP BY 1
)
SELECT ps.patientunitstayid, ps.hospitalid, ps.ethnicity, ps.gender, SAFE_CAST(ps.age AS FLOAT64) AS age,
    ps.hidden_flag,
    CASE WHEN ps.discharge_status = 'Expired' THEN 1 ELSE 0 END AS died,
    s.max_creatinine, s.max_bilirubin, s.min_platelets, s.max_lactate
FROM per_stay ps
LEFT JOIN severity s ON ps.patientunitstayid = s.patientunitstayid
WHERE s.max_lactate < 2.0;

-- F3: First-48h + normal lactate
WITH paired AS (
    SELECT v.patientunitstayid, pt.hospitaldischargestatus, v.observationoffset,
        CASE WHEN SAFE_CAST(v.sao2 AS FLOAT64) >= 94 AND SAFE_CAST(l.labresult AS FLOAT64) < 88 THEN 1 ELSE 0 END AS hidden_severe
    FROM `physionet-data.eicu_crd.vitalperiodic` v
    JOIN `physionet-data.eicu_crd.patient` pt ON v.patientunitstayid = pt.patientunitstayid
    JOIN `physionet-data.eicu_crd.lab` l ON l.patientunitstayid = v.patientunitstayid
        AND l.labname = 'O2 Sat (%)'
        AND ABS(l.labresultoffset - v.observationoffset) <= 60
    WHERE v.sao2 IS NOT NULL
        AND SAFE_CAST(v.sao2 AS FLOAT64) BETWEEN 50 AND 100
        AND SAFE_CAST(l.labresult AS FLOAT64) BETWEEN 50 AND 100
        AND v.observationoffset <= 2880
),
per_stay AS (
    SELECT patientunitstayid,
        ANY_VALUE(hospitaldischargestatus) AS discharge_status,
        CASE WHEN SUM(hidden_severe) > 0 THEN 'Hidden (48h + normal lactate)' ELSE 'No hidden' END AS hidden_group
    FROM paired
    GROUP BY 1
),
lactate AS (
    SELECT patientunitstayid, MAX(SAFE_CAST(labresult AS FLOAT64)) AS max_lactate
    FROM `physionet-data.eicu_crd.lab`
    WHERE labname = 'lactate' AND SAFE_CAST(labresult AS FLOAT64) BETWEEN 0.1 AND 50
    GROUP BY 1
)
SELECT ps.hidden_group,
    COUNT(*) AS n_stays,
    ROUND(100.0 * COUNTIF(ps.discharge_status = 'Expired') / COUNT(*), 1) AS mortality_pct
FROM per_stay ps
JOIN lactate la ON ps.patientunitstayid = la.patientunitstayid
WHERE la.max_lactate < 2.0
GROUP BY 1
ORDER BY 1;


-- =============================================================================
-- PART G: GOLDEN SUBSET (MIMIC-III WAVEFORM) — Manuscript Section 2.9, Table 9
-- =============================================================================
-- Requires: algorithm_benchmarks.mimic3_waveform_subjects (from S2 pipeline or BQ).
-- G1: Master demographics + outcomes. G2: SpO2/SaO2 paired. G3–G6: Summaries.

-- G1: Golden clinical master
CREATE OR REPLACE TABLE `candor-health-486219.algorithm_benchmarks.golden_clinical_master` AS
WITH waveform_subjects AS (
    SELECT subject_id FROM `candor-health-486219.algorithm_benchmarks.mimic3_waveform_subjects`
),
first_admission AS (
    SELECT a.SUBJECT_ID, a.HADM_ID, a.ETHNICITY, a.ADMISSION_TYPE, a.DIAGNOSIS,
        a.HOSPITAL_EXPIRE_FLAG, a.ADMITTIME, a.DISCHTIME,
        DATETIME_DIFF(a.DISCHTIME, a.ADMITTIME, HOUR) AS los_hours,
        ROW_NUMBER() OVER (PARTITION BY a.SUBJECT_ID ORDER BY a.ADMITTIME) AS rn
    FROM `physionet-data.mimiciii_clinical.admissions` a
    JOIN waveform_subjects w ON a.SUBJECT_ID = w.subject_id
)
SELECT fa.SUBJECT_ID AS subject_id, p.GENDER AS gender, fa.ETHNICITY AS ethnicity,
    CASE WHEN fa.ETHNICITY LIKE '%BLACK%' OR fa.ETHNICITY LIKE '%AFRICAN%' THEN 'Black'
         WHEN fa.ETHNICITY LIKE '%WHITE%' THEN 'White'
         WHEN fa.ETHNICITY LIKE '%HISPANIC%' OR fa.ETHNICITY LIKE '%LATINO%' THEN 'Hispanic'
         WHEN fa.ETHNICITY LIKE '%ASIAN%' THEN 'Asian' ELSE 'Other' END AS race_group,
    DATETIME_DIFF(fa.ADMITTIME, p.DOB, YEAR) AS approx_age,
    fa.HADM_ID AS hadm_id, fa.ADMISSION_TYPE AS admission_type, fa.DIAGNOSIS AS primary_diagnosis,
    fa.HOSPITAL_EXPIRE_FLAG AS died, fa.los_hours,
    p.DOD IS NOT NULL AS died_any, p.EXPIRE_FLAG AS expire_flag
FROM first_admission fa
JOIN `physionet-data.mimiciii_clinical.patients` p ON fa.SUBJECT_ID = p.SUBJECT_ID
WHERE fa.rn = 1;

-- G2: Golden SpO2/SaO2 paired (MIMIC-III itemids 646,220277 SpO2; 834,220227 SaO2)
CREATE OR REPLACE TABLE `candor-health-486219.algorithm_benchmarks.golden_spo2_paired` AS
WITH waveform_subjects AS (
    SELECT subject_id FROM `candor-health-486219.algorithm_benchmarks.mimic3_waveform_subjects`
),
spo2 AS (
    SELECT c.SUBJECT_ID, c.HADM_ID, c.CHARTTIME, c.VALUENUM AS spo2
    FROM `physionet-data.mimiciii_clinical.chartevents` c
    JOIN waveform_subjects w ON c.SUBJECT_ID = w.subject_id
    WHERE c.ITEMID IN (646, 220277) AND c.VALUENUM IS NOT NULL AND c.VALUENUM BETWEEN 50 AND 100
),
sao2 AS (
    SELECT c.SUBJECT_ID, c.HADM_ID, c.CHARTTIME, c.VALUENUM AS sao2
    FROM `physionet-data.mimiciii_clinical.chartevents` c
    JOIN waveform_subjects w ON c.SUBJECT_ID = w.subject_id
    WHERE c.ITEMID IN (834, 220227) AND c.VALUENUM IS NOT NULL AND c.VALUENUM BETWEEN 50 AND 100
),
paired AS (
    SELECT s.SUBJECT_ID, s.HADM_ID, s.CHARTTIME AS spo2_time, s.spo2, a.CHARTTIME AS sao2_time, a.sao2,
        s.spo2 - a.sao2 AS bias,
        ABS(DATETIME_DIFF(s.CHARTTIME, a.CHARTTIME, MINUTE)) AS time_diff_min,
        CASE WHEN s.spo2 >= 94 AND a.sao2 < 88 THEN 1 ELSE 0 END AS hidden_severe,
        CASE WHEN s.spo2 >= 92 AND a.sao2 < 88 THEN 1 ELSE 0 END AS hidden_moderate,
        ROW_NUMBER() OVER (PARTITION BY s.SUBJECT_ID, s.HADM_ID, s.CHARTTIME ORDER BY ABS(DATETIME_DIFF(s.CHARTTIME, a.CHARTTIME, MINUTE))) AS rn
    FROM spo2 s
    JOIN sao2 a ON s.SUBJECT_ID = a.SUBJECT_ID AND s.HADM_ID = a.HADM_ID
        AND ABS(DATETIME_DIFF(s.CHARTTIME, a.CHARTTIME, MINUTE)) <= 30
)
SELECT SUBJECT_ID AS subject_id, HADM_ID AS hadm_id, spo2_time, spo2, sao2_time, sao2, bias, time_diff_min, hidden_severe, hidden_moderate
FROM paired
WHERE rn = 1;

-- G3: Hidden hypoxemia by race (waveform cohort)
SELECT m.race_group,
    CASE WHEN SUM(p.hidden_severe) > 0 THEN 'Had hidden hypoxemia' ELSE 'No hidden hypoxemia' END AS hidden_group,
    COUNT(DISTINCT m.subject_id) AS n_subjects, SUM(m.died) AS deaths,
    ROUND(100.0 * SUM(m.died) / COUNT(DISTINCT m.subject_id), 1) AS mortality_pct,
    ROUND(AVG(p.bias), 3) AS avg_bias, SUM(p.hidden_severe) AS total_hidden_events
FROM `candor-health-486219.algorithm_benchmarks.golden_clinical_master` m
LEFT JOIN (SELECT subject_id, bias, hidden_severe FROM `candor-health-486219.algorithm_benchmarks.golden_spo2_paired`) p ON m.subject_id = p.subject_id
GROUP BY 1, 2
ORDER BY 1, 2;

-- G4: Vital signs summary (waveform subjects)
SELECT m.race_group, m.gender, COUNT(DISTINCT m.subject_id) AS n_subjects,
    ROUND(AVG(m.approx_age), 1) AS avg_age, ROUND(AVG(m.los_hours), 1) AS avg_los_hours,
    SUM(m.died) AS deaths, ROUND(100.0 * SUM(m.died) / COUNT(DISTINCT m.subject_id), 1) AS mortality_pct
FROM `candor-health-486219.algorithm_benchmarks.golden_clinical_master` m
GROUP BY 1, 2
ORDER BY 1, 2;

-- G5: SOFA-like severity (labs) for waveform subjects
SELECT m.race_group, m.died, COUNT(DISTINCT m.subject_id) AS n,
    ROUND(AVG(cr.creatinine), 2) AS avg_creatinine, ROUND(AVG(bi.bilirubin), 2) AS avg_bilirubin,
    ROUND(AVG(pl.platelets), 0) AS avg_platelets, ROUND(AVG(la.lactate), 2) AS avg_lactate
FROM `candor-health-486219.algorithm_benchmarks.golden_clinical_master` m
LEFT JOIN (SELECT SUBJECT_ID, HADM_ID, AVG(VALUENUM) AS creatinine FROM `physionet-data.mimiciii_clinical.labevents` WHERE ITEMID IN (50912) AND VALUENUM BETWEEN 0.1 AND 30 GROUP BY 1, 2) cr ON m.subject_id = cr.SUBJECT_ID AND m.hadm_id = cr.HADM_ID
LEFT JOIN (SELECT SUBJECT_ID, HADM_ID, AVG(VALUENUM) AS bilirubin FROM `physionet-data.mimiciii_clinical.labevents` WHERE ITEMID IN (50885) AND VALUENUM BETWEEN 0 AND 100 GROUP BY 1, 2) bi ON m.subject_id = bi.SUBJECT_ID AND m.hadm_id = bi.HADM_ID
LEFT JOIN (SELECT SUBJECT_ID, HADM_ID, AVG(VALUENUM) AS platelets FROM `physionet-data.mimiciii_clinical.labevents` WHERE ITEMID IN (51265) AND VALUENUM BETWEEN 1 AND 2000 GROUP BY 1, 2) pl ON m.subject_id = pl.SUBJECT_ID AND m.hadm_id = pl.HADM_ID
LEFT JOIN (SELECT SUBJECT_ID, HADM_ID, AVG(VALUENUM) AS lactate FROM `physionet-data.mimiciii_clinical.labevents` WHERE ITEMID IN (50813) AND VALUENUM BETWEEN 0.1 AND 50 GROUP BY 1, 2) la ON m.subject_id = la.SUBJECT_ID AND m.hadm_id = la.HADM_ID
GROUP BY 1, 2
ORDER BY 1, 2;

-- G6: Top diagnoses (waveform subjects)
SELECT m.race_group, d.ICD9_CODE, di.SHORT_TITLE, COUNT(*) AS n
FROM `candor-health-486219.algorithm_benchmarks.golden_clinical_master` m
JOIN `physionet-data.mimiciii_clinical.diagnoses_icd` d ON m.subject_id = d.SUBJECT_ID AND m.hadm_id = d.HADM_ID
LEFT JOIN `physionet-data.mimiciii_clinical.d_icd_diagnoses` di ON d.ICD9_CODE = di.ICD9_CODE
GROUP BY 1, 2, 3
HAVING n >= 20
ORDER BY 1, n DESC;


-- =============================================================================
-- PART H: PRIMARY MORTALITY CROSS-TABS (Manuscript Tables 4 and 5)
-- =============================================================================
-- These produce the core hidden-hypoxemia-vs-mortality tables that are the
-- central finding of the paper. Tables 4 (MIMIC-IV) and 5 (eICU-CRD).

-- H1: MIMIC-IV — Hidden hypoxemia × mortality by race (Table 4)
WITH spo2 AS (
    SELECT SUBJECT_ID, HADM_ID, CHARTTIME, VALUENUM AS spo2
    FROM `physionet-data.mimiciv_3_1_icu.chartevents`
    WHERE ITEMID = 220277 AND VALUENUM BETWEEN 50 AND 100
),
sao2 AS (
    SELECT SUBJECT_ID, HADM_ID, CHARTTIME, VALUENUM AS sao2
    FROM `physionet-data.mimiciv_3_1_icu.chartevents`
    WHERE ITEMID = 220227 AND VALUENUM BETWEEN 50 AND 100
),
paired AS (
    SELECT s.SUBJECT_ID, s.HADM_ID, s.spo2, a.sao2,
        CASE WHEN s.spo2 >= 94 AND a.sao2 < 88 THEN 1 ELSE 0 END AS hidden_severe,
        ROW_NUMBER() OVER (PARTITION BY s.SUBJECT_ID, s.HADM_ID, s.CHARTTIME
            ORDER BY ABS(DATETIME_DIFF(s.CHARTTIME, a.CHARTTIME, MINUTE))) AS rn
    FROM spo2 s
    JOIN sao2 a ON s.SUBJECT_ID = a.SUBJECT_ID AND s.HADM_ID = a.HADM_ID
        AND ABS(DATETIME_DIFF(s.CHARTTIME, a.CHARTTIME, MINUTE)) <= 30
),
per_stay AS (
    SELECT SUBJECT_ID, HADM_ID,
        CASE WHEN SUM(hidden_severe) > 0 THEN 'Yes' ELSE 'No' END AS hidden_group
    FROM paired WHERE rn = 1
    GROUP BY 1, 2
)
SELECT adm.race,
    ps.hidden_group,
    COUNT(*) AS n_stays,
    SUM(adm.hospital_expire_flag) AS deaths,
    ROUND(100.0 * SUM(adm.hospital_expire_flag) / COUNT(*), 1) AS mortality_pct
FROM per_stay ps
JOIN `physionet-data.mimiciv_3_1_hosp.admissions` adm
    ON ps.SUBJECT_ID = adm.subject_id AND ps.HADM_ID = adm.hadm_id
GROUP BY 1, 2
ORDER BY 1, 2;

-- H2: eICU-CRD — Hidden hypoxemia × mortality by ethnicity (Table 5)
WITH paired AS (
    SELECT v.patientunitstayid, pt.ethnicity, pt.hospitaldischargestatus,
        CASE WHEN SAFE_CAST(v.sao2 AS FLOAT64) >= 94
              AND SAFE_CAST(l.labresult AS FLOAT64) < 88 THEN 1 ELSE 0 END AS hidden_severe
    FROM `physionet-data.eicu_crd.vitalperiodic` v
    JOIN `physionet-data.eicu_crd.patient` pt ON v.patientunitstayid = pt.patientunitstayid
    JOIN `physionet-data.eicu_crd.lab` l ON l.patientunitstayid = v.patientunitstayid
        AND l.labname = 'O2 Sat (%)'
        AND ABS(l.labresultoffset - v.observationoffset) <= 60
    WHERE v.sao2 IS NOT NULL
        AND SAFE_CAST(v.sao2 AS FLOAT64) BETWEEN 50 AND 100
        AND SAFE_CAST(l.labresult AS FLOAT64) BETWEEN 50 AND 100
),
per_stay AS (
    SELECT patientunitstayid,
        ANY_VALUE(ethnicity) AS ethnicity,
        ANY_VALUE(hospitaldischargestatus) AS discharge_status,
        SUM(hidden_severe) AS hidden_count,
        CASE WHEN SUM(hidden_severe) > 0 THEN 'Yes' ELSE 'No' END AS hidden_group
    FROM paired GROUP BY 1
)
SELECT ethnicity, hidden_group,
    COUNT(*) AS n_stays,
    ROUND(100.0 * COUNTIF(discharge_status = 'Expired') / COUNT(*), 1) AS mortality_pct,
    ROUND(AVG(hidden_count), 1) AS avg_hidden_events
FROM per_stay
GROUP BY 1, 2
ORDER BY 1, 2;


-- =============================================================================
-- PART I: SEVERITY MARKERS (Manuscript Table 6, Panels A and B)
-- =============================================================================

-- I1: MIMIC-IV severity markers by hidden status and race (Table 6, Panel A)
WITH spo2 AS (
    SELECT SUBJECT_ID, HADM_ID, CHARTTIME, VALUENUM AS spo2
    FROM `physionet-data.mimiciv_3_1_icu.chartevents`
    WHERE ITEMID = 220277 AND VALUENUM BETWEEN 50 AND 100
),
sao2 AS (
    SELECT SUBJECT_ID, HADM_ID, CHARTTIME, VALUENUM AS sao2
    FROM `physionet-data.mimiciv_3_1_icu.chartevents`
    WHERE ITEMID = 220227 AND VALUENUM BETWEEN 50 AND 100
),
paired AS (
    SELECT s.SUBJECT_ID, s.HADM_ID,
        CASE WHEN s.spo2 >= 94 AND a.sao2 < 88 THEN 1 ELSE 0 END AS hidden_severe,
        ROW_NUMBER() OVER (PARTITION BY s.SUBJECT_ID, s.HADM_ID, s.CHARTTIME
            ORDER BY ABS(DATETIME_DIFF(s.CHARTTIME, a.CHARTTIME, MINUTE))) AS rn
    FROM spo2 s
    JOIN sao2 a ON s.SUBJECT_ID = a.SUBJECT_ID AND s.HADM_ID = a.HADM_ID
        AND ABS(DATETIME_DIFF(s.CHARTTIME, a.CHARTTIME, MINUTE)) <= 30
),
per_stay AS (
    SELECT SUBJECT_ID, HADM_ID,
        CASE WHEN SUM(hidden_severe) > 0 THEN 'Had hidden hypoxemia' ELSE 'No hidden hypoxemia' END AS hidden_group
    FROM paired WHERE rn = 1 GROUP BY 1, 2
),
severity AS (
    SELECT SUBJECT_ID, HADM_ID,
        MAX(CASE WHEN ITEMID = 50912 THEN VALUENUM END) AS max_creatinine,
        MAX(CASE WHEN ITEMID = 50885 THEN VALUENUM END) AS max_bilirubin,
        MIN(CASE WHEN ITEMID = 51265 THEN VALUENUM END) AS min_platelets,
        MAX(CASE WHEN ITEMID = 50813 THEN VALUENUM END) AS max_lactate
    FROM `physionet-data.mimiciv_3_1_hosp.labevents`
    WHERE ITEMID IN (50912, 50885, 51265, 50813) AND VALUENUM IS NOT NULL
    GROUP BY 1, 2
)
SELECT adm.race, ps.hidden_group, COUNT(*) AS n,
    ROUND(100.0 * SUM(adm.hospital_expire_flag) / COUNT(*), 1) AS mortality_pct,
    ROUND(AVG(s.max_creatinine), 2) AS avg_creatinine,
    ROUND(AVG(s.max_bilirubin), 2) AS avg_bilirubin,
    ROUND(AVG(s.min_platelets), 0) AS avg_min_platelets,
    ROUND(AVG(s.max_lactate), 2) AS avg_lactate
FROM per_stay ps
JOIN `physionet-data.mimiciv_3_1_hosp.admissions` adm ON ps.SUBJECT_ID = adm.subject_id AND ps.HADM_ID = adm.hadm_id
LEFT JOIN severity s ON ps.SUBJECT_ID = s.SUBJECT_ID AND ps.HADM_ID = s.HADM_ID
WHERE adm.race IN ('WHITE', 'BLACK/AFRICAN AMERICAN', 'ASIAN', 'OTHER')
GROUP BY 1, 2 ORDER BY 1, 2;

-- I2: eICU severity markers by hidden status and ethnicity (Table 6, Panel B)
-- Uses the same per-stay + severity structure as Part B but outputs the cross-tab.
WITH paired AS (
    SELECT v.patientunitstayid, pt.ethnicity, pt.hospitaldischargestatus,
        CASE WHEN SAFE_CAST(v.sao2 AS FLOAT64) >= 94 AND SAFE_CAST(l.labresult AS FLOAT64) < 88 THEN 1 ELSE 0 END AS hidden_severe
    FROM `physionet-data.eicu_crd.vitalperiodic` v
    JOIN `physionet-data.eicu_crd.patient` pt ON v.patientunitstayid = pt.patientunitstayid
    JOIN `physionet-data.eicu_crd.lab` l ON l.patientunitstayid = v.patientunitstayid
        AND l.labname = 'O2 Sat (%)' AND ABS(l.labresultoffset - v.observationoffset) <= 60
    WHERE v.sao2 IS NOT NULL AND SAFE_CAST(v.sao2 AS FLOAT64) BETWEEN 50 AND 100 AND SAFE_CAST(l.labresult AS FLOAT64) BETWEEN 50 AND 100
),
per_stay AS (
    SELECT patientunitstayid, ANY_VALUE(ethnicity) AS ethnicity, ANY_VALUE(hospitaldischargestatus) AS discharge_status,
        CASE WHEN SUM(hidden_severe) > 0 THEN 'Had hidden hypoxemia' ELSE 'No hidden hypoxemia' END AS hidden_group
    FROM paired GROUP BY 1
),
severity AS (
    SELECT patientunitstayid,
        MAX(CASE WHEN labname = 'creatinine' THEN SAFE_CAST(labresult AS FLOAT64) END) AS max_creatinine,
        MAX(CASE WHEN labname = 'total bilirubin' THEN SAFE_CAST(labresult AS FLOAT64) END) AS max_bilirubin,
        MIN(CASE WHEN labname = 'platelets x 1000' THEN SAFE_CAST(labresult AS FLOAT64) END) AS min_platelets,
        MAX(CASE WHEN labname = 'lactate' THEN SAFE_CAST(labresult AS FLOAT64) END) AS max_lactate
    FROM `physionet-data.eicu_crd.lab` GROUP BY 1
)
SELECT ps.ethnicity, ps.hidden_group, COUNT(*) AS n_stays,
    ROUND(100.0 * COUNTIF(ps.discharge_status = 'Expired') / COUNT(*), 1) AS mortality_pct,
    ROUND(AVG(s.max_creatinine), 2) AS avg_creatinine,
    ROUND(AVG(s.max_bilirubin), 2) AS avg_bilirubin,
    ROUND(AVG(s.min_platelets), 0) AS avg_min_platelets,
    ROUND(AVG(s.max_lactate), 2) AS avg_lactate
FROM per_stay ps
LEFT JOIN severity s ON ps.patientunitstayid = s.patientunitstayid
GROUP BY 1, 2 ORDER BY 1, 2;


-- =============================================================================
-- PART J: OPEN OXIMETRY REPOSITORY — Mechanistic Queries
--         (Manuscript Tables 2 Panel A reading-level, 3, 7, 8, Section 3.4.3)
-- =============================================================================
-- These use the internal dataset: candor-health-486219.algorithm_benchmarks
-- Replace the project/dataset prefix if running under a different account.
-- Open Oximetry tables: pulseox, bloodgas, spectro (joined on encounter_id).

-- J1: Reading-level bias by melanin bucket (Table 3)
SELECT
    CASE
        WHEN sp.melanin_index < 1.0 THEN 'Low (<1.0)'
        WHEN sp.melanin_index < 1.5 THEN 'Medium (1.0-1.5)'
        WHEN sp.melanin_index < 2.0 THEN 'High (1.5-2.0)'
        ELSE 'Very High (>2.0)'
    END AS melanin_level,
    COUNT(*) AS n,
    ROUND(AVG(p.saturation - b.so2), 3) AS mean_bias
FROM `candor-health-486219.algorithm_benchmarks.pulseox` p
JOIN `candor-health-486219.algorithm_benchmarks.bloodgas` b
    ON p.encounter_id = b.encounter_id AND p.sample_number = b.sample_number
JOIN `candor-health-486219.algorithm_benchmarks.spectro` sp
    ON p.encounter_id = sp.encounter_id
WHERE p.saturation BETWEEN 50 AND 100
    AND b.so2 BETWEEN 50 AND 100
GROUP BY 1
ORDER BY 1;

-- J2: Bias by perfusion index × melanin level (Table 7)
SELECT
    CASE
        WHEN p.pi < 0.5 THEN 'Very Low (<0.5)'
        WHEN p.pi < 1.0 THEN 'Low (0.5-1.0)'
        WHEN p.pi < 2.0 THEN 'Normal (1.0-2.0)'
        WHEN p.pi < 5.0 THEN 'Good (2.0-5.0)'
        ELSE 'High (5.0+)'
    END AS pi_level,
    CASE
        WHEN sp.melanin_index < 1.0 THEN 'Low Melanin (<1.0)'
        WHEN sp.melanin_index < 1.5 THEN 'Mid Melanin (1.0-1.5)'
        ELSE 'High Melanin (>=1.5)'
    END AS melanin_group,
    COUNT(*) AS n,
    ROUND(AVG(p.saturation - b.so2), 3) AS mean_bias
FROM `candor-health-486219.algorithm_benchmarks.pulseox` p
JOIN `candor-health-486219.algorithm_benchmarks.bloodgas` b
    ON p.encounter_id = b.encounter_id AND p.sample_number = b.sample_number
JOIN `candor-health-486219.algorithm_benchmarks.spectro` sp
    ON p.encounter_id = sp.encounter_id
WHERE p.saturation BETWEEN 50 AND 100
    AND b.so2 BETWEEN 50 AND 100
    AND p.pi IS NOT NULL
GROUP BY 1, 2
ORDER BY 1, 2;

-- J3: Triple interaction PI × melanin × severe hypoxia (Table 8)
SELECT
    CASE
        WHEN p.pi < 0.5 THEN 'Very Low (<0.5)'
        WHEN p.pi < 1.0 THEN 'Low (0.5-1.0)'
        ELSE 'Normal+ (>=1.0)'
    END AS pi_level,
    CASE WHEN sp.melanin_index >= 1.5 THEN 'High (>=1.5)' ELSE 'Low (<1.0)' END AS melanin_group,
    COUNT(*) AS n,
    ROUND(AVG(p.saturation - b.so2), 2) AS mean_bias,
    SUM(CASE WHEN p.saturation >= 94 AND b.so2 < 88 THEN 1 ELSE 0 END) AS hidden_severe_events,
    ROUND(100.0 * SUM(CASE WHEN p.saturation >= 94 AND b.so2 < 88 THEN 1 ELSE 0 END) / COUNT(*), 1) AS pct_hidden_severe
FROM `candor-health-486219.algorithm_benchmarks.pulseox` p
JOIN `candor-health-486219.algorithm_benchmarks.bloodgas` b
    ON p.encounter_id = b.encounter_id AND p.sample_number = b.sample_number
JOIN `candor-health-486219.algorithm_benchmarks.spectro` sp
    ON p.encounter_id = sp.encounter_id
WHERE p.saturation BETWEEN 50 AND 100
    AND b.so2 BETWEEN 50 AND 100
    AND b.so2 < 88  -- severe hypoxia only
    AND p.pi IS NOT NULL
    AND sp.melanin_index IS NOT NULL
GROUP BY 1, 2
ORDER BY 1, 2;

-- J4: "Device displays safe" — true SaO2 when SpO2 reads 93-95% (Section 3.4.3)
SELECT
    ROUND(sp.melanin_index, 1) AS melanin_bucket,
    COUNT(*) AS n_readings_at_93_95,
    ROUND(AVG(b.so2), 1) AS avg_true_sao2,
    SUM(CASE WHEN b.so2 < 88 THEN 1 ELSE 0 END) AS count_actually_severe,
    ROUND(100.0 * SUM(CASE WHEN b.so2 < 88 THEN 1 ELSE 0 END) / COUNT(*), 1) AS pct_actually_severe
FROM `candor-health-486219.algorithm_benchmarks.pulseox` p
JOIN `candor-health-486219.algorithm_benchmarks.bloodgas` b
    ON p.encounter_id = b.encounter_id AND p.sample_number = b.sample_number
JOIN `candor-health-486219.algorithm_benchmarks.spectro` sp
    ON p.encounter_id = sp.encounter_id
WHERE p.saturation BETWEEN 93 AND 95
    AND b.so2 BETWEEN 50 AND 100
GROUP BY 1
ORDER BY 1;

-- J5: Reading-level bias by eICU ethnicity (Table 2 Panel A — reading-level)
SELECT
    pt.ethnicity,
    COUNT(*) AS n_paired_readings,
    ROUND(AVG(SAFE_CAST(v.sao2 AS FLOAT64) - SAFE_CAST(l.labresult AS FLOAT64)), 3) AS mean_bias
FROM `physionet-data.eicu_crd.vitalperiodic` v
JOIN `physionet-data.eicu_crd.patient` pt ON v.patientunitstayid = pt.patientunitstayid
JOIN `physionet-data.eicu_crd.lab` l ON l.patientunitstayid = v.patientunitstayid
    AND l.labname = 'O2 Sat (%)'
    AND ABS(l.labresultoffset - v.observationoffset) <= 60
WHERE v.sao2 IS NOT NULL
    AND SAFE_CAST(v.sao2 AS FLOAT64) BETWEEN 50 AND 100
    AND SAFE_CAST(l.labresult AS FLOAT64) BETWEEN 50 AND 100
GROUP BY 1
ORDER BY 1;

-- J6: MIMIC-IV bias by race (Section 3.2.3 — includes +3.40% Black subgroup)
WITH spo2 AS (
    SELECT SUBJECT_ID, HADM_ID, CHARTTIME, VALUENUM AS spo2
    FROM `physionet-data.mimiciv_3_1_icu.chartevents`
    WHERE ITEMID = 220277 AND VALUENUM BETWEEN 50 AND 100
),
sao2 AS (
    SELECT SUBJECT_ID, HADM_ID, CHARTTIME, VALUENUM AS sao2
    FROM `physionet-data.mimiciv_3_1_icu.chartevents`
    WHERE ITEMID = 220227 AND VALUENUM BETWEEN 50 AND 100
),
paired AS (
    SELECT s.SUBJECT_ID, s.HADM_ID, s.spo2, a.sao2, s.spo2 - a.sao2 AS bias,
        ROW_NUMBER() OVER (PARTITION BY s.SUBJECT_ID, s.HADM_ID, s.CHARTTIME
            ORDER BY ABS(DATETIME_DIFF(s.CHARTTIME, a.CHARTTIME, MINUTE))) AS rn
    FROM spo2 s
    JOIN sao2 a ON s.SUBJECT_ID = a.SUBJECT_ID AND s.HADM_ID = a.HADM_ID
        AND ABS(DATETIME_DIFF(s.CHARTTIME, a.CHARTTIME, MINUTE)) <= 30
)
SELECT adm.race,
    COUNT(*) AS n_paired_readings,
    ROUND(AVG(p.bias), 3) AS mean_bias
FROM paired p
JOIN `physionet-data.mimiciv_3_1_hosp.admissions` adm
    ON p.SUBJECT_ID = adm.subject_id AND p.HADM_ID = adm.hadm_id
WHERE p.rn = 1
GROUP BY 1
ORDER BY 1;
```
