## Supplementary material for "Perfusion-Dependent Melanin Bias in Pulse Oximetry and ICU Mortality Across 209 U.S. Hospitals: A Multicenter Retrospective Analysis of 52 Million Readings": Gehring_2026_PulseOx_Supplements.zip: Supplement_S2_Waveform_Extraction.py

```
#!/usr/bin/env python3
"""
Golden Subset Pipeline — Candor Systems
========================================
Run on Vertex AI JupyterLab (where GCS access is native).

This script does THREE things:
  Phase 1: INVENTORY  — scan all 883 Black + comparison subjects' waveform headers,
                         identify which have PLETH (PPG), ECG, ABP, SpO2 numerics.
  Phase 2: EXTRACT    — for subjects with PLETH, read the waveform segments and
                         compute signal-quality features (AC/DC, perfusion proxy, SNR).
  Phase 3: OUTPUT     — write features to CSV → upload to BigQuery for joining
                         with clinical outcomes.

Usage:
  # Phase 1 only (fast, ~30 min for all subjects)
  python golden_subset_pipeline.py --phase inventory

  # Phase 2 (heavy, run on subset first)
  python golden_subset_pipeline.py --phase extract --max-subjects 50

  # Phase 3 (upload results)
  python golden_subset_pipeline.py --phase upload

Requirements (install on Vertex AI):
  pip install wfdb numpy pandas google-cloud-storage google-cloud-bigquery tqdm
"""

import argparse
import csv
import io
import json
import os
import re
import struct
import subprocess
import sys
import tempfile
import time
from pathlib import Path

try:
    import numpy as np
    import pandas as pd
    from tqdm import tqdm
except ImportError:
    print("Missing deps. Run: pip install numpy pandas tqdm")
    sys.exit(1)

# ---------------------------------------------------------------------------
# CONFIG
# ---------------------------------------------------------------------------
GCS_BUCKET = "gs://candor-biosignal-data-lake"
WAVEFORM_ROOT = f"{GCS_BUCKET}/mimic3wdb"
MATCHED_ROOT = f"{WAVEFORM_ROOT}/matched"

BQ_PROJECT = "candor-health-486219"
BQ_DATASET = "algorithm_benchmarks"
BQ_SUBJECTS_TABLE = f"{BQ_PROJECT}.{BQ_DATASET}.mimic3_waveform_subjects"

OUTPUT_DIR = "/tmp/golden_subset"
INVENTORY_CSV = f"{OUTPUT_DIR}/waveform_inventory.csv"
FEATURES_CSV = f"{OUTPUT_DIR}/waveform_features.csv"

# Target ethnicities for the Golden Subset
TARGET_ETHNICITIES = [
    "BLACK/AFRICAN AMERICAN",
    "HISPANIC OR LATINO",
    "ASIAN",
]
# Comparison group
COMPARISON_ETHNICITY = "WHITE"
COMPARISON_SAMPLE_N = 1000  # random sample of White subjects for comparison


# ---------------------------------------------------------------------------
# HELPERS
# ---------------------------------------------------------------------------

def run_cmd(cmd, capture=True):
    """Run a shell command, return stdout."""
    result = subprocess.run(cmd, shell=True, capture_output=capture, text=True)
    if result.returncode != 0 and capture:
        # Don't fail hard — some gsutil commands return non-zero for empty results
        pass
    return result.stdout.strip() if capture else ""


def gsutil_cat(path):
    """Read a GCS file to string."""
    return run_cmd(f"gsutil cat {path}")


def gsutil_ls(path):
    """List a GCS path, return list of paths."""
    out = run_cmd(f"gsutil ls {path}")
    return [line.strip() for line in out.split("\n") if line.strip()]


def parse_hea_header(header_text):
    """
    Parse a WFDB .hea file (text content) to extract:
      - record name, n_signals, sample_rate, n_samples
      - signal names
      - segment list (for multi-segment records)
    """
    lines = [l.strip() for l in header_text.strip().split("\n") if l.strip() and not l.startswith("#")]
    if not lines:
        return None

    # First line: record_name n_sig fs n_samp [base_time base_date]
    first = lines[0].split()
    info = {
        "record_name": first[0].split("/")[0],
        "n_signals": 0,
        "fs": 0,
        "n_samples": 0,
        "signals": [],
        "segments": [],
    }

    # Multi-segment records have format: record/n_segments n_sig fs n_samp
    if "/" in first[0]:
        parts = first[0].split("/")
        info["record_name"] = parts[0]
        info["n_segments"] = int(parts[1]) if len(parts) > 1 else 0
        info["n_signals"] = int(first[1]) if len(first) > 1 else 0
        info["fs"] = int(first[2]) if len(first) > 2 else 0
        info["n_samples"] = int(first[3]) if len(first) > 3 else 0

        # Remaining lines are segment entries: segment_name n_samples [~ gap_samples]
        for line in lines[1:]:
            seg_parts = line.split()
            seg_name = seg_parts[0]
            seg_len = int(seg_parts[1]) if len(seg_parts) > 1 else 0
            info["segments"].append({"name": seg_name, "n_samples": seg_len})
    else:
        info["n_signals"] = int(first[1]) if len(first) > 1 else 0
        info["fs"] = int(first[2]) if len(first) > 2 else 0
        info["n_samples"] = int(first[3]) if len(first) > 3 else 0

        # Remaining lines are signal specs: filename format gain baseline units resolution ...  sig_name
        for line in lines[1:]:
            parts = line.split()
            if len(parts) >= 9:
                info["signals"].append(parts[-1])  # last field = signal name
            elif len(parts) >= 2:
                info["signals"].append(parts[-1])

    return info


def parse_layout_header(header_text):
    """
    Parse a _layout.hea file to get the full signal list for multi-segment records.
    Returns list of signal names.
    """
    lines = [l.strip() for l in header_text.strip().split("\n") if l.strip() and not l.startswith("#")]
    if not lines:
        return []

    signals = []
    for line in lines[1:]:  # skip first line (record info)
        parts = line.split()
        if len(parts) >= 2:
            signals.append(parts[-1])  # last field = signal name
    return signals


# ---------------------------------------------------------------------------
# PHASE 1: INVENTORY
# ---------------------------------------------------------------------------

def get_subject_ids_from_bq(ethnicities=None):
    """
    Query BigQuery for subject IDs with waveforms, filtered by ethnicity.
    Returns dict: {subject_id: {"ethnicity": ..., "hospital_expire_flag": ...}}
    """
    if ethnicities is None:
        ethnicities = TARGET_ETHNICITIES + [COMPARISON_ETHNICITY]

    eth_list = ", ".join([f"'{e}'" for e in ethnicities])

    query = f"""
    SELECT DISTINCT
        w.subject_id,
        a.ETHNICITY,
        a.HOSPITAL_EXPIRE_FLAG,
        p.GENDER,
        DATETIME_DIFF(a.ADMITTIME, p.DOB, YEAR) AS approx_age
    FROM `{BQ_SUBJECTS_TABLE}` w
    JOIN `physionet-data.mimiciii_clinical.admissions` a
        ON w.subject_id = a.SUBJECT_ID
    JOIN `physionet-data.mimiciii_clinical.patients` p
        ON w.subject_id = p.SUBJECT_ID
    WHERE a.ETHNICITY IN ({eth_list})
    ORDER BY a.ETHNICITY, w.subject_id
    """

    print(f"Querying BigQuery for subjects with ethnicities: {ethnicities}...")
    # Write query to temp file to avoid shell quoting issues (ethnicity strings contain single quotes)
    with tempfile.NamedTemporaryFile(mode="w", suffix=".sql", delete=False) as f:
        f.write(query.strip())
        qpath = f.name
    try:
        with open(qpath, "r") as f:
            proc = subprocess.run(
                ["bq", "query", "--nouse_legacy_sql", "--format=json", "--max_rows=50000"],
                stdin=f,
                capture_output=True,
                text=True,
            )
        result = proc.stdout if proc.returncode == 0 else (proc.stderr or proc.stdout or "")
    finally:
        os.unlink(qpath)

    if not result:
        print("ERROR: BigQuery returned empty result. Check access.")
        return {}

    try:
        rows = json.loads(result)
    except json.JSONDecodeError:
        print(f"ERROR parsing BQ result. First 500 chars: {result[:500]}")
        return {}

    subjects = {}
    for row in rows:
        sid = int(row["subject_id"])
        if sid not in subjects:
            subjects[sid] = {
                "ethnicity": row.get("ETHNICITY", "UNKNOWN"),
                "died": row.get("HOSPITAL_EXPIRE_FLAG", "0") == "1",
                "gender": row.get("GENDER", ""),
                "approx_age": row.get("approx_age", ""),
            }

    # Sample comparison group if too large
    target_sids = {sid for sid, info in subjects.items() if info["ethnicity"] != COMPARISON_ETHNICITY}
    comparison_sids = {sid for sid, info in subjects.items() if info["ethnicity"] == COMPARISON_ETHNICITY}

    if len(comparison_sids) > COMPARISON_SAMPLE_N:
        import random
        random.seed(42)
        comparison_sids = set(random.sample(list(comparison_sids), COMPARISON_SAMPLE_N))
        subjects = {sid: info for sid, info in subjects.items()
                    if sid in target_sids or sid in comparison_sids}

    print(f"  Total subjects: {len(subjects)}")
    for eth in ethnicities:
        n = sum(1 for s in subjects.values() if s["ethnicity"] == eth)
        print(f"    {eth}: {n}")

    return subjects


def subject_id_to_gcs_path(subject_id):
    """
    Convert subject_id → GCS matched path.
    e.g. subject_id 109 → matched/p00/p000109/
    """
    padded = f"p{subject_id:06d}"
    prefix = padded[:3]  # p00, p01, etc.
    return f"{MATCHED_ROOT}/{prefix}/{padded}/"


def inventory_subject(subject_id):
    """
    Read the master .hea for a subject's waveform record to find:
    - which signals are present (PLETH, ECG, ABP, etc.)
    - total duration
    - sample rate
    - whether numerics exist

    Returns dict with inventory info, or None if no waveform found.
    """
    gcs_path = subject_id_to_gcs_path(subject_id)

    # List the subject's folder
    files = gsutil_ls(gcs_path)
    if not files:
        return None

    # Find the master .hea file (not _layout, not segment _XXXX)
    hea_files = [f for f in files if f.endswith(".hea")]
    master_hea = None
    layout_hea = None
    numeric_hea = None

    for f in hea_files:
        basename = f.split("/")[-1]
        if "_layout" in basename:
            layout_hea = f
        elif "n.hea" in basename:
            numeric_hea = f
        elif re.match(r'^p\d+\.hea$', basename) or re.match(r'^\d+\.hea$', basename):
            # Could be master — check if it's not a segment
            if not re.match(r'.*_\d{4}\.hea$', basename):
                master_hea = f

    if not master_hea and not layout_hea:
        # Fallback: just take the shortest-named .hea
        non_segment = [f for f in hea_files if not re.match(r'.*_\d{4}\.hea$', f.split("/")[-1])
                       and "_layout" not in f and "n.hea" not in f]
        if non_segment:
            master_hea = sorted(non_segment, key=len)[0]

    # Parse master header
    signals = []
    fs = 0
    n_samples = 0
    n_segments = 0
    duration_hours = 0

    if master_hea:
        header_text = gsutil_cat(master_hea)
        if header_text:
            info = parse_hea_header(header_text)
            if info:
                fs = info.get("fs", 0)
                n_samples = info.get("n_samples", 0)
                n_segments = len(info.get("segments", []))
                if fs > 0 and n_samples > 0:
                    duration_hours = round(n_samples / fs / 3600, 2)

    # Parse layout for signal names
    if layout_hea:
        layout_text = gsutil_cat(layout_hea)
        if layout_text:
            signals = parse_layout_header(layout_text)

    # Parse numerics header
    has_numerics = False
    numeric_signals = []
    if numeric_hea:
        has_numerics = True
        num_text = gsutil_cat(numeric_hea)
        if num_text:
            numeric_signals = parse_layout_header(num_text)
            if not numeric_signals:
                # Try parsing as regular header
                num_info = parse_hea_header(num_text)
                if num_info:
                    numeric_signals = num_info.get("signals", [])

    # Classify signals
    has_pleth = any("pleth" in s.lower() or "ppg" in s.lower() for s in signals)
    has_ecg = any(s.upper() in ["I", "II", "III", "V", "AVR", "AVL", "AVF", "MCL", "ECG"]
                  or "ecg" in s.lower() for s in signals)
    has_abp = any("abp" in s.lower() for s in signals)
    has_spo2_numeric = any("spo2" in s.lower() or "sp02" in s.lower() for s in numeric_signals)

    # Count total files (dat + hea)
    n_files = len(files)

    return {
        "subject_id": subject_id,
        "gcs_path": gcs_path,
        "n_files": n_files,
        "fs": fs,
        "n_samples": n_samples,
        "n_segments": n_segments,
        "duration_hours": duration_hours,
        "signals": signals,
        "has_pleth": has_pleth,
        "has_ecg": has_ecg,
        "has_abp": has_abp,
        "has_numerics": has_numerics,
        "has_spo2_numeric": has_spo2_numeric,
        "numeric_signals": numeric_signals,
    }


def run_inventory(subjects):
    """Phase 1: Inventory all subjects' waveforms."""
    os.makedirs(OUTPUT_DIR, exist_ok=True)

    print(f"\n{'='*70}")
    print(f"PHASE 1: WAVEFORM INVENTORY — {len(subjects)} subjects")
    print(f"{'='*70}\n")

    results = []
    errors = []

    for sid in tqdm(sorted(subjects.keys()), desc="Scanning waveforms"):
        try:
            inv = inventory_subject(sid)
            if inv:
                inv["ethnicity"] = subjects[sid]["ethnicity"]
                inv["died"] = subjects[sid]["died"]
                inv["gender"] = subjects[sid]["gender"]
                results.append(inv)
            else:
                errors.append({"subject_id": sid, "error": "no waveform found"})
        except Exception as e:
            errors.append({"subject_id": sid, "error": str(e)[:100]})

    # Write inventory CSV
    if results:
        df = pd.DataFrame(results)
        df.to_csv(INVENTORY_CSV, index=False)
        print(f"\nInventory saved: {INVENTORY_CSV}")

        # Summary
        print(f"\n{'='*70}")
        print("INVENTORY SUMMARY")
        print(f"{'='*70}")
        print(f"Total subjects with waveforms: {len(results)}")
        print(f"Subjects with NO waveform:     {len(errors)}")
        print()

        for eth in df["ethnicity"].unique():
            sub = df[df["ethnicity"] == eth]
            print(f"  {eth}:")
            print(f"    Total:     {len(sub)}")
            print(f"    Has PLETH: {sub['has_pleth'].sum()} ({100*sub['has_pleth'].mean():.1f}%)")
            print(f"    Has ECG:   {sub['has_ecg'].sum()} ({100*sub['has_ecg'].mean():.1f}%)")
            print(f"    Has ABP:   {sub['has_abp'].sum()} ({100*sub['has_abp'].mean():.1f}%)")
            print(f"    Has SpO2 numeric: {sub['has_spo2_numeric'].sum()} ({100*sub['has_spo2_numeric'].mean():.1f}%)")
            print(f"    Deaths:    {sub['died'].sum()}")
            print(f"    Avg duration: {sub['duration_hours'].mean():.1f} hours")
            print()

        # The key number
        pleth_subjects = df[df["has_pleth"]]
        print(f"\n*** GOLDEN SUBSET CANDIDATES (has PLETH + demographics): {len(pleth_subjects)} ***")
        for eth in pleth_subjects["ethnicity"].unique():
            n = len(pleth_subjects[pleth_subjects["ethnicity"] == eth])
            print(f"    {eth}: {n}")

    if errors:
        err_path = f"{OUTPUT_DIR}/inventory_errors.csv"
        pd.DataFrame(errors).to_csv(err_path, index=False)
        print(f"\nErrors saved: {err_path}")

    return results


# ---------------------------------------------------------------------------
# PHASE 2: EXTRACT FEATURES (from subjects with PLETH)
# ---------------------------------------------------------------------------

def download_segment(gcs_path, segment_name, local_dir):
    """Download a single segment (.hea + .dat) to local dir."""
    os.makedirs(local_dir, exist_ok=True)
    for ext in [".hea", ".dat"]:
        src = f"{gcs_path}{segment_name}{ext}"
        dst = os.path.join(local_dir, f"{segment_name}{ext}")
        run_cmd(f"gsutil -q cp {src} {dst}", capture=False)
    return os.path.join(local_dir, segment_name)


def extract_ppg_features(signal, fs=125):
    """
    Extract signal quality features from a PPG (PLETH) segment.

    Returns dict with:
      - mean_amplitude: mean of signal
      - ac_component: std of signal (proxy for AC)
      - dc_component: mean of signal (proxy for DC)
      - ac_dc_ratio: AC/DC ratio (perfusion proxy)
      - snr_db: signal-to-noise ratio estimate
      - signal_entropy: Shannon entropy of amplitude distribution
      - zero_crossing_rate: rate of zero crossings (quality indicator)
      - pct_flat: % of samples that are flat/saturated
      - pct_valid: % of non-NaN, non-zero samples
    """
    if signal is None or len(signal) == 0:
        return None

    # Clean
    signal = np.array(signal, dtype=float)
    valid_mask = np.isfinite(signal) & (signal != 0)
    pct_valid = valid_mask.mean() * 100

    if pct_valid < 10:
        return {"pct_valid": pct_valid, "quality": "unusable"}

    clean = signal[valid_mask]

    # Basic stats
    dc = np.mean(clean)
    ac = np.std(clean)
    ac_dc = ac / abs(dc) if abs(dc) > 1e-6 else 0

    # SNR estimate: power of signal / power of high-freq noise
    # Simple approach: SNR ≈ variance(bandpass 0.5-5Hz) / variance(>10Hz)
    try:
        from scipy.signal import butter, filtfilt

        # Bandpass 0.5-5 Hz (physiological PPG)
        b_bp, a_bp = butter(3, [0.5, 5.0], btype='bandpass', fs=fs)
        ppg_filtered = filtfilt(b_bp, a_bp, clean)

        # Highpass >10 Hz (noise)
        b_hp, a_hp = butter(3, 10.0, btype='highpass', fs=fs)
        noise = filtfilt(b_hp, a_hp, clean)

        sig_power = np.var(ppg_filtered)
        noise_power = np.var(noise)
        snr_db = 10 * np.log10(sig_power / noise_power) if noise_power > 1e-12 else 40.0
    except ImportError:
        # scipy not available — rough estimate
        snr_db = None

    # Zero crossing rate
    centered = clean - np.mean(clean)
    zero_crossings = np.sum(np.diff(np.sign(centered)) != 0)
    zcr = zero_crossings / len(centered)

    # Flatness (saturated signal)
    diffs = np.diff(clean)
    pct_flat = (np.sum(np.abs(diffs) < 1e-6) / len(diffs)) * 100

    # Entropy (amplitude distribution)
    hist, _ = np.histogram(clean, bins=50, density=True)
    hist = hist[hist > 0]
    entropy = -np.sum(hist * np.log2(hist)) if len(hist) > 0 else 0

    return {
        "mean_amplitude": round(float(dc), 4),
        "ac_component": round(float(ac), 4),
        "dc_component": round(float(dc), 4),
        "ac_dc_ratio": round(float(ac_dc), 6),
        "snr_db": round(float(snr_db), 2) if snr_db is not None else None,
        "signal_entropy": round(float(entropy), 4),
        "zero_crossing_rate": round(float(zcr), 6),
        "pct_flat": round(float(pct_flat), 2),
        "pct_valid": round(float(pct_valid), 2),
        "quality": "good" if (pct_valid > 80 and pct_flat < 50) else "poor",
    }


def extract_features_for_subject(subject_id, gcs_path, max_segments=5):
    """
    Download up to max_segments PLETH segments for a subject,
    extract PPG features from each, return aggregated features.
    """
    try:
        import wfdb
    except ImportError:
        print("WFDB not installed. Run: pip install wfdb")
        return None

    local_dir = os.path.join(OUTPUT_DIR, "waveforms", f"p{subject_id:06d}")
    os.makedirs(local_dir, exist_ok=True)

    # Get master header
    padded = f"p{subject_id:06d}"
    master_hea_path = f"{gcs_path}{padded}.hea"
    # Also try numeric record name format
    files = gsutil_ls(gcs_path)
    hea_files = [f for f in files if f.endswith(".hea") and "_layout" not in f and "n.hea" not in f]

    # Find layout to know which signal index is PLETH
    layout_files = [f for f in files if "_layout.hea" in f]
    pleth_idx = None
    if layout_files:
        layout_text = gsutil_cat(layout_files[0])
        layout_signals = parse_layout_header(layout_text)
        for i, sig in enumerate(layout_signals):
            if "pleth" in sig.lower() or "ppg" in sig.lower():
                pleth_idx = i
                break

    if pleth_idx is None:
        return None

    # Find segment .hea files (e.g. 3000003_0001.hea)
    segment_files = sorted([f for f in hea_files if re.match(r'.*_\d{4}\.hea$', f.split("/")[-1])])

    if not segment_files:
        return None

    # Process up to max_segments
    all_features = []
    segments_processed = 0

    for seg_file in segment_files[:max_segments * 3]:  # try more in case some fail
        if segments_processed >= max_segments:
            break

        seg_name = seg_file.split("/")[-1].replace(".hea", "")

        try:
            # Download segment
            for ext in [".hea", ".dat"]:
                src = f"{gcs_path}{seg_name}{ext}"
                dst = os.path.join(local_dir, f"{seg_name}{ext}")
                if not os.path.exists(dst):
                    run_cmd(f"gsutil -q cp {src} {dst}", capture=False)

            # Read with wfdb
            record = wfdb.rdrecord(os.path.join(local_dir, seg_name))

            if pleth_idx < record.p_signal.shape[1]:
                ppg_signal = record.p_signal[:, pleth_idx]
                features = extract_ppg_features(ppg_signal, fs=record.fs)
                if features and features.get("quality") != "unusable":
                    features["segment"] = seg_name
                    features["n_samples"] = len(ppg_signal)
                    features["duration_sec"] = len(ppg_signal) / record.fs
                    all_features.append(features)
                    segments_processed += 1

        except Exception as e:
            continue  # skip bad segments

    if not all_features:
        return None

    # Aggregate across segments
    good_features = [f for f in all_features if f.get("quality") == "good"]
    use_features = good_features if good_features else all_features

    agg = {
        "subject_id": subject_id,
        "n_segments_analyzed": len(all_features),
        "n_good_segments": len(good_features),
        "total_duration_sec": sum(f["duration_sec"] for f in all_features),
        "mean_ac_dc_ratio": np.mean([f["ac_dc_ratio"] for f in use_features]),
        "mean_snr_db": np.mean([f["snr_db"] for f in use_features if f.get("snr_db") is not None]),
        "mean_pct_valid": np.mean([f["pct_valid"] for f in use_features]),
        "mean_pct_flat": np.mean([f["pct_flat"] for f in use_features]),
        "mean_entropy": np.mean([f["signal_entropy"] for f in use_features]),
        "mean_zcr": np.mean([f["zero_crossing_rate"] for f in use_features]),
        "overall_quality": "good" if len(good_features) > len(all_features) / 2 else "poor",
    }

    return agg


def run_extract(inventory_df, max_subjects=None):
    """Phase 2: Extract PPG features for subjects with PLETH."""
    pleth_df = inventory_df[inventory_df["has_pleth"] == True].copy()

    if max_subjects:
        pleth_df = pleth_df.head(max_subjects)

    print(f"\n{'='*70}")
    print(f"PHASE 2: PPG FEATURE EXTRACTION — {len(pleth_df)} subjects")
    print(f"{'='*70}\n")

    results = []
    for _, row in tqdm(pleth_df.iterrows(), total=len(pleth_df), desc="Extracting features"):
        sid = int(row["subject_id"])
        gcs_path = row["gcs_path"]

        features = extract_features_for_subject(sid, gcs_path, max_segments=5)
        if features:
            features["ethnicity"] = row["ethnicity"]
            features["died"] = row["died"]
            features["gender"] = row.get("gender", "")
            results.append(features)

    if results:
        df = pd.DataFrame(results)
        df.to_csv(FEATURES_CSV, index=False)
        print(f"\nFeatures saved: {FEATURES_CSV}")

        # Summary by ethnicity
        print(f"\n{'='*70}")
        print("FEATURE EXTRACTION SUMMARY")
        print(f"{'='*70}")

        for eth in df["ethnicity"].unique():
            sub = df[df["ethnicity"] == eth]
            print(f"\n  {eth} (n={len(sub)}):")
            print(f"    Mean AC/DC ratio:  {sub['mean_ac_dc_ratio'].mean():.6f}")
            if sub["mean_snr_db"].notna().any():
                print(f"    Mean SNR (dB):     {sub['mean_snr_db'].mean():.2f}")
            print(f"    Mean % valid:      {sub['mean_pct_valid'].mean():.1f}%")
            print(f"    Good quality:      {(sub['overall_quality']=='good').sum()} / {len(sub)}")

        # The key comparison
        print(f"\n{'='*70}")
        print("KEY COMPARISON: Signal Quality by Ethnicity")
        print(f"{'='*70}")

        summary = df.groupby("ethnicity").agg({
            "mean_ac_dc_ratio": ["mean", "std"],
            "mean_snr_db": "mean",
            "mean_pct_valid": "mean",
            "overall_quality": lambda x: (x == "good").mean() * 100,
        }).round(4)
        print(summary.to_string())

        # Mortality comparison
        if df["died"].any():
            print(f"\n{'='*70}")
            print("Signal Quality vs Mortality")
            print(f"{'='*70}")
            mort = df.groupby("died").agg({
                "mean_ac_dc_ratio": "mean",
                "mean_snr_db": "mean",
                "mean_pct_valid": "mean",
            }).round(4)
            print(mort.to_string())

    return results


# ---------------------------------------------------------------------------
# PHASE 3: UPLOAD TO BIGQUERY
# ---------------------------------------------------------------------------

def run_upload():
    """Phase 3: Upload features CSV to BigQuery."""
    print(f"\n{'='*70}")
    print("PHASE 3: UPLOAD TO BIGQUERY")
    print(f"{'='*70}\n")

    # Upload inventory
    if os.path.exists(INVENTORY_CSV):
        table = f"{BQ_PROJECT}:{BQ_DATASET}.golden_subset_inventory"
        cmd = (f"bq load --autodetect --replace "
               f"--source_format=CSV {table} {INVENTORY_CSV}")
        print(f"Uploading inventory → {table}")
        run_cmd(cmd, capture=False)
        print("  Done.")

    # Upload features
    if os.path.exists(FEATURES_CSV):
        table = f"{BQ_PROJECT}:{BQ_DATASET}.golden_subset_features"
        cmd = (f"bq load --autodetect --replace "
               f"--source_format=CSV {table} {FEATURES_CSV}")
        print(f"Uploading features → {table}")
        run_cmd(cmd, capture=False)
        print("  Done.")

    print("\nTables available for joining:")
    print(f"  {BQ_PROJECT}.{BQ_DATASET}.golden_subset_inventory")
    print(f"  {BQ_PROJECT}.{BQ_DATASET}.golden_subset_features")
    print("\nJoin with clinical data:")
    print(f"""
    SELECT
        f.subject_id, f.ethnicity, f.died,
        f.mean_ac_dc_ratio, f.mean_snr_db, f.overall_quality,
        a.HOSPITAL_EXPIRE_FLAG, a.DIAGNOSIS
    FROM `{BQ_PROJECT}.{BQ_DATASET}.golden_subset_features` f
    JOIN `physionet-data.mimiciii_clinical.admissions` a
        ON f.subject_id = a.SUBJECT_ID
    ORDER BY f.ethnicity, f.mean_ac_dc_ratio
    """)


# ---------------------------------------------------------------------------
# MAIN
# ---------------------------------------------------------------------------

def main():
    parser = argparse.ArgumentParser(description="Candor Golden Subset Pipeline")
    parser.add_argument("--phase", choices=["inventory", "extract", "upload", "all"],
                        default="inventory", help="Which phase to run")
    parser.add_argument("--max-subjects", type=int, default=None,
                        help="Limit number of subjects (for testing)")
    parser.add_argument("--inventory-csv", type=str, default=INVENTORY_CSV,
                        help="Path to existing inventory CSV (skip Phase 1)")

    args = parser.parse_args()

    os.makedirs(OUTPUT_DIR, exist_ok=True)

    if args.phase in ("inventory", "all"):
        subjects = get_subject_ids_from_bq()
        if not subjects:
            print("No subjects found. Check BigQuery access.")
            sys.exit(1)
        inventory = run_inventory(subjects)

    if args.phase in ("extract", "all"):
        # Load inventory
        if os.path.exists(args.inventory_csv):
            print(f"Loading inventory from {args.inventory_csv}")
            inv_df = pd.read_csv(args.inventory_csv)
        elif args.phase == "all" and inventory:
            inv_df = pd.DataFrame(inventory)
        else:
            print(f"No inventory found at {args.inventory_csv}. Run --phase inventory first.")
            sys.exit(1)

        run_extract(inv_df, max_subjects=args.max_subjects)

    if args.phase in ("upload", "all"):
        run_upload()


if __name__ == "__main__":
    main()
```
