## Supplementary material for "Perfusion-Dependent Melanin Bias in Pulse Oximetry and ICU Mortality Across 209 U.S. Hospitals: A Multicenter Retrospective Analysis of 52 Million Readings": Gehring_2026_PulseOx_Supplements.zip: Supplement_S3_Sensitivity_Analyses.md

# Supplement S3 — Sensitivity Analyses

**Manuscript:** Gehring 2026. Perfusion-Dependent Melanin Bias in Pulse Oximetry and ICU Mortality Across 209 U.S. Hospitals.

Results from sensitivity analyses (Section 2.10). Queries: Supplement S1, Part C.

---

## Table S3.1 — Bias by ethnicity with 95% CI (eICU; Table 2 source)

Patient-level mean bias per stay, then group statistics. **Manuscript Table 2.**

| Ethnicity         | N stays | Mean bias (%) | SD   | 95% CI lower | 95% CI upper |
|-------------------|--------|---------------|------|--------------|--------------|
| (blank)           | 618    | 0.76          | 3.93 | 0.45         | 1.07         |
| African American  | 5,458  | 2.19          | 5.88 | 2.04         | 2.35         |
| Asian             | 869    | 2.08          | 5.10 | 1.74         | 2.42         |
| Caucasian         | 42,853 | 1.39          | 4.65 | 1.35         | 1.44         |
| Hispanic          | 2,487  | 1.53          | 3.93 | 1.38         | 1.68         |
| Native American   | 439    | 1.81          | 4.47 | 1.39         | 2.23         |
| Other/Unknown     | 2,561  | 1.72          | 4.42 | 1.55         | 1.89         |

---

## Table S3.2 — Pairing window 15 min (MIMIC-IV)

Mortality by hidden hypoxemia when SpO2/SaO2 paired within **15 minutes** (vs. primary 30 min). **Manuscript Section 3.3.5.**

| Race                      | Hidden group | N stays | Deaths | Mortality (%) |
|---------------------------|--------------|--------|--------|---------------|
| BLACK/AFRICAN AMERICAN    | Hidden       | 51     | 14     | 27.5          |
| BLACK/AFRICAN AMERICAN    | No hidden    | 983    | 227    | 23.1          |
| WHITE                     | Hidden       | 305    | 89     | 29.2          |
| WHITE                     | No hidden    | 9,647  | 1,590  | 16.5          |

---

## Table S3.3 — Alternate threshold (SpO2 >= 92%, SaO2 < 90%)

Hidden hypoxemia defined with relaxed thresholds. **Manuscript Section 3.3.5.**

| Ethnicity        | Hidden group   | N stays | Mortality (%) |
|------------------|----------------|--------|---------------|
| (blank)          | Hidden (92/90) | 126    | 19.8          |
| (blank)          | No hidden      | 492    | 11.8          |
| African American | Hidden (92/90) | 1,189  | 26.1          |
| African American | No hidden      | 4,269  | 14.5          |
| Asian            | Hidden (92/90) | 176    | 33.0          |
| Asian            | No hidden      | 693    | 14.4          |
| Caucasian        | Hidden (92/90) | 8,324  | 29.4          |
| Caucasian        | No hidden      | 34,529 | 14.6          |
| Hispanic         | Hidden (92/90) | 444    | 33.3          |
| Hispanic         | No hidden      | 2,043  | 16.5          |
| Native American  | Hidden (92/90) | 85     | 42.4          |
| Native American  | No hidden      | 354    | 13.3          |
| Other/Unknown    | Hidden (92/90) | 525    | 27.6          |
| Other/Unknown    | No hidden      | 2,036  | 14.3          |

---

## Table S3.4 — Patient-level aggregated bias (eICU)

Bias averaged within each patient (one value per stay). **Manuscript Section 3.3.5** — racial gradient preserved (African American median +1.49% vs Caucasian +0.92%).

| Ethnicity        | N stays | Mean bias (%) | SD   | Median bias (%) |
|------------------|--------|---------------|------|-----------------|
| (blank)          | 618    | 0.76          | 3.93 | 0.42            |
| African American | 5,458  | 2.19          | 5.88 | 1.49            |
| Asian            | 869    | 2.08          | 5.10 | 1.27            |
| Caucasian        | 42,853 | 1.39          | 4.65 | 0.92            |
| Hispanic         | 2,487  | 1.53          | 3.93 | 1.26            |
| Native American  | 439    | 1.81          | 4.47 | 1.47            |
| Other/Unknown    | 2,561  | 1.72          | 4.42 | 1.10            |

---

## Table S3.5 — Minimum 3 paired readings per stay (eICU)

Restriction to stays with at least 3 paired SpO2/SaO2 readings. **Manuscript Section 3.3.5** — approximately 2x mortality ratio virtually unchanged.

| Ethnicity        | Hidden group | N stays | Mortality (%) |
|------------------|--------------|--------|---------------|
| (blank)          | Hidden       | 79     | 25.3          |
| (blank)          | No hidden    | 529    | 11.3          |
| African American | Hidden       | 937    | 26.6          |
| African American | No hidden    | 4,439  | 14.9          |
| Asian            | Hidden       | 133    | 34.6          |
| Asian            | No hidden    | 724    | 15.2          |
| Caucasian        | Hidden       | 5,727  | 31.1          |
| Caucasian        | No hidden    | 36,512 | 15.4          |
| Hispanic         | Hidden       | 318    | 34.9          |
| Hispanic         | No hidden    | 2,132  | 17.4          |
| Native American  | Hidden       | 53     | 39.6          |
| Native American  | No hidden    | 381    | 16.0          |
| Other/Unknown    | Hidden       | 382    | 30.9          |
| Other/Unknown    | No hidden    | 2,149  | 14.7          |

---

All analyses use Standard SQL in BigQuery; see Supplement S1 for exact queries.
