## Supplementary material for "Perfusion-Dependent Melanin Bias in Pulse Oximetry and ICU Mortality Across 209 U.S. Hospitals: A Multicenter Retrospective Analysis of 52 Million Readings": Gehring_2026_PulseOx_Supplements.zip: Supplement_S4_Missingness_Analysis.md

# Supplement S4 — Missingness Analysis

**Manuscript:** Gehring 2026. Perfusion-Dependent Melanin Bias in Pulse Oximetry and ICU Mortality Across 209 U.S. Hospitals.

Demographics of ICU stays **included** vs **excluded** from the mortality analysis (eICU-CRD). Stays were included if they had ≥1 paired SpO₂/SaO₂ reading within 60 minutes. **Manuscript Section 2.11, Supplement S4.**

Query: Supplement S1, Part D (Query D1).

---

## Table S4.1 — Included vs excluded stays by ethnicity (eICU-CRD)

| Group    | Ethnicity        | N stays | Mean age | % Male | Mortality (%) |
|----------|------------------|--------|----------|--------|---------------|
| Excluded | (blank)          | 1,672  | 60.9     | 53.3   | 6.7           |
| Excluded | African American | 15,850 | 55.4     | 50.7   | 5.9           |
| Excluded | Asian            | 2,401  | 62.2     | 51.3   | 6.3           |
| Excluded | Caucasian        | 112,432| 63.0     | 53.7   | 5.7           |
| Excluded | Hispanic         | 4,977  | 57.7     | 54.1   | 5.0           |
| Excluded | Native American  | 1,261  | 52.4     | 49.3   | 4.4           |
| Excluded | Other/Unknown    | 6,981  | 57.6     | 56.3   | 5.6           |
| Included | (blank)          | 618    | 63.2     | 57.4   | 13.4          |
| Included | African American | 5,458  | 58.8     | 52.5   | 17.0          |
| Included | Asian            | 869    | 63.0     | 55.2   | 18.2          |
| Included | Caucasian        | 42,853 | 64.3     | 55.6   | 17.4          |
| Included | Hispanic         | 2,487  | 63.1     | 53.4   | 19.5          |
| Included | Native American  | 439    | 52.0     | 52.8   | 18.9          |
| Included | Other/Unknown    | 2,561  | 60.4     | 59.1   | 17.1          |

**Interpretation:** Included stays (those with paired SpO₂/SaO₂) are older on average, slightly more often male, and have substantially higher mortality than excluded stays — consistent with sicker patients receiving more frequent ABGs and monitoring. This selection is acknowledged in the manuscript limitations; the mortality analysis applies to the cohort with sufficient oximetry–ABG pairing for classification.

---

*Query: Supplement S1, Part D.*
