## Supplementary material for "Perfusion-Dependent Melanin Bias in Pulse Oximetry and ICU Mortality Across 209 U.S. Hospitals: A Multicenter Retrospective Analysis of 52 Million Readings": Gehring_2026_PulseOx_Supplements.zip: Supplement_S5_Full_Regression_Model.md

# Supplement S5 — Full Multivariable Logistic Regression Model

**Manuscript:** Gehring 2026. Perfusion-Dependent Melanin Bias in Pulse Oximetry and ICU Mortality Across 209 U.S. Hospitals.

Logistic regression for in-hospital mortality with cluster-robust standard errors by hospital (eICU-CRD). **Manuscript Section 2.7, Table 6c.**

---

## Model specification

- **Outcome:** In-hospital death (binary).
- **Primary exposure:** Hidden hypoxemia (binary; ≥1 paired reading with SpO₂ ≥ 94% and SaO₂ < 88%).
- **Covariates:** Age (continuous), sex (categorical), race/ethnicity (categorical), peak creatinine, peak bilirubin, nadir platelets, peak lactate.
- **Cluster-robust SEs:** By `hospitalid` to account for within-hospital correlation.
- **Software:** statsmodels (Python) `logit(..., cov_type='cluster', cov_kwds={'groups': hospitalid})`.

Regression input table: Supplement S1, Part B (Query B1). Export `eicu_regression_input` to CSV, drop rows with missing key covariates, then fit.

---

## Full cohort — coefficients and odds ratios

**N (after listwise deletion):** 24,009 stays. **Pseudo R²:** 0.161. **LLR p-value:** < 0.001.

| Variable                 | Coef    | Std err | z    | P > \|z\| | [0.025  | 0.975] | Odds ratio |
|--------------------------|--------|---------|------|-----------|---------|--------|------------|
| Intercept                | -4.464 | 0.204   | -21.9| 0.000     | -4.863  | -4.065 | 0.0115     |
| C(gender)[T.Male]        | 0.037  | 0.044   | 0.84 | 0.401     | -0.049  | 0.123  | 1.038      |
| C(ethnicity)[T.Asian]    | -0.030 | 0.148   | -0.20| 0.838     | -0.320  | 0.260  | 0.970      |
| C(ethnicity)[T.Caucasian]| 0.276  | 0.102   | 2.72 | 0.007     | 0.077   | 0.475  | 1.318      |
| C(ethnicity)[T.Hispanic] | 0.221  | 0.139   | 1.58 | 0.113     | -0.053  | 0.494  | 1.247      |
| **hidden_flag**          | **0.619** | **0.049** | **12.68** | **0.000** | **0.524** | **0.715** | **1.857** |
| age                      | 0.029  | 0.002   | 18.17| 0.000     | 0.026   | 0.033  | 1.030      |
| max_creatinine           | 0.051  | 0.008   | 6.23 | 0.000     | 0.035   | 0.067  | 1.053      |
| max_bilirubin            | 0.054  | 0.008   | 7.10 | 0.000     | 0.039   | 0.069  | 1.055      |
| min_platelets            | -0.001 | 0.000   | -2.54| 0.011     | -0.001  | -0.000 | 0.999      |
| max_lactate              | 0.198  | 0.006   | 32.25| 0.000     | 0.186   | 0.210  | 1.219      |

**Primary result (full cohort):** Adjusted OR for hidden hypoxemia **1.86** (95% CI 1.69–2.04), p < 0.001.

---

## Normal-lactate subgroup (max lactate < 2.0 mmol/L)

**Purpose:** Test whether the mortality association is independent of circulatory collapse (Section 2.7.3, Table 6h, Section 3.3.7).

**N:** 9,951 stays. **Pseudo R²:** 0.055. **LLR p-value:** < 0.001.

| Variable                 | Coef    | Std err | z    | P > \|z\| | [0.025  | 0.975] | Odds ratio | 95% CI OR   |
|--------------------------|--------|---------|------|-----------|---------|--------|------------|-------------|
| Intercept                | -4.564 | 0.352   | -13.0| 0.000     | -5.255  | -3.874 | 0.0104     | 0.005–0.021 |
| C(gender)[T.Male]        | 0.138  | 0.070   | 1.99 | 0.047     | 0.002   | 0.274  | 1.148      | 1.00–1.32   |
| C(ethnicity)[T.Asian]    | -0.200 | 0.328   | -0.61| 0.542     | -0.843  | 0.443  | 0.819      | 0.43–1.56   |
| C(ethnicity)[T.Caucasian]| 0.174  | 0.154   | 1.13 | 0.258     | -0.128  | 0.477  | 1.191      | 0.88–1.61   |
| C(ethnicity)[T.Hispanic] | 0.128  | 0.236   | 0.54 | 0.588     | -0.335  | 0.590  | 1.136      | 0.72–1.80   |
| **hidden_flag**          | **0.624** | **0.074** | **8.39** | **0.000** | **0.478** | **0.770** | **1.87** | **1.61–2.16** |
| age                      | 0.036  | 0.003   | 10.90| 0.000     | 0.029   | 0.042  | 1.036      | 1.03–1.04   |
| max_creatinine           | 0.042  | 0.013   | 3.13 | 0.002     | 0.016   | 0.068  | 1.043      | 1.02–1.07   |
| max_bilirubin            | 0.069  | 0.015   | 4.71 | 0.000     | 0.040   | 0.098  | 1.071      | 1.04–1.10   |
| min_platelets            | -0.002 | 0.001   | -3.19| 0.001     | -0.003  | -0.001 | 0.998      | 0.997–0.999 |

**Primary result (normal lactate):** Adjusted OR for hidden hypoxemia **1.87** (95% CI 1.61–2.16), p < 0.001.

---

*Input tables: S1 Part B (full cohort), S1 Part F2 (normal-lactate table).*
