## Supplementary material for "Perfusion-Dependent Melanin Bias in Pulse Oximetry and ICU Mortality Across 209 U.S. Hospitals: A Multicenter Retrospective Analysis of 52 Million Readings": Gehring_2026_PulseOx_Supplements.zip: Supplement_S6_Reproducibility_Guide.md

# Supplement S6 — Dataset Access and Reproducibility Guide

**Manuscript:** Gehring 2026. Perfusion-Dependent Melanin Bias in Pulse Oximetry and ICU Mortality Across 209 U.S. Hospitals.

This guide describes how to obtain access to the underlying data and reproduce the analyses.

---

## 1. Data sources and access

All clinical data used in this study are from **PhysioNet** (https://physionet.org) and require **credentialed access** (free after completing training and signing the relevant data use agreements).

| Dataset | PhysioNet project | BigQuery location | Role in manuscript |
|---------|-------------------|-------------------|--------------------|
| MIMIC-IV v3.1 | MIMIC-IV | `physionet-data.mimiciv_3_1_hosp`, `physionet-data.mimiciv_3_1_icu` | Single-center mortality, bias (Table 2), sensitivity (15-min pairing) |
| eICU-CRD | eICU Collaborative Research Database | `physionet-data.eicu_crd` | National mortality (55,178 stays), regression, landmark, lactate |
| Open Oximetry Repository | openoximetry | (internal copy; see PhysioNet) | Perfusion × melanin mechanism (52.4M readings) |
| MIMIC-III Clinical | MIMIC-III Clinical Database | `physionet-data.mimiciii_clinical` | Golden Subset clinical linkage |
| MIMIC-III Waveform | MIMIC-III Waveform Database | N/A (files on GCS or local) | PPG signal quality (Section 2.9, Table 9) |

**Important:** MIMIC-IV v3.1 dataset names use **no "v"**: `mimiciv_3_1_hosp` and `mimiciv_3_1_icu` (not `mimiciv_v3_1_*`).

### Obtaining BigQuery access

1. Create a **Google Cloud** account and a project (or use an existing one).
2. Request access to each PhysioNet project that hosts BigQuery data (MIMIC-IV, eICU, MIMIC-III, Open Oximetry as applicable).
3. Complete the required training (e.g., CITI) and sign the data use agreement for each project.
4. After approval, add your Google account to the PhysioNet project’s BigQuery access list. PhysioNet documentation specifies the exact steps (e.g., “Request access” → “GCP BigQuery”).
5. In Google Cloud Console, set your billing project. You can query `physionet-data` from your own project; you will be charged for query bytes processed by your project.

---

## 2. Reproducing the SQL analyses (S1)

1. **Open BigQuery** in Google Cloud Console (console.cloud.google.com/bigquery).
2. Ensure the account with PhysioNet access is selected and the correct billing project is set.
3. Open **Supplement S1** (`Supplement_S1_SQL_Queries.sql`).
4. Run each query block in order. Some queries **create tables** in a destination dataset (e.g. `candor-health-486219.algorithm_benchmarks`). To run those, you must either:
   - Create a dataset in your own project and replace `candor-health-486219.algorithm_benchmarks` with your project and dataset, or  
   - Omit the CREATE TABLE queries and run only SELECT queries if you do not need to persist intermediate tables.
5. **Parts A–F** use only `physionet-data` (MIMIC-IV, eICU). **Part G** (Golden Subset) also requires a table `mimic3_waveform_subjects` in your project; that table is produced by the waveform pipeline (Section 3 below) or by building the subject list from the MIMIC-III waveform `matched/` folder and uploading to BigQuery.

Results of each SELECT can be exported to CSV for comparison with the manuscript and Supplements S3–S5.

---

## 3. Reproducing the waveform pipeline (S2)

The waveform signal-quality analysis (Section 2.9, Table 9) uses the **MIMIC-III Waveform Database** and the script in **Supplement S2** (`Supplement_S2_Waveform_Extraction.py`).

### Prerequisites

- **MIMIC-III Waveform Database** downloaded (or accessible) in a structure compatible with the script (e.g. `matched/p00/p000020/` with `.hea` and `.dat` files). The manuscript used waveforms on **Google Cloud Storage** (e.g. `gs://candor-biosignal-data-lake/mimic3wdb/`).
- **Python 3** with: `numpy`, `pandas`, `tqdm`; for Phase 2 (extract): `wfdb`, `scipy`. On Vertex AI: `pip install numpy pandas tqdm wfdb scipy google-cloud-storage google-cloud-bigquery`.
- **BigQuery** table of waveform subject IDs (`mimic3_waveform_subjects`) if the script is configured to pull the cohort from BigQuery.
- **Google Cloud authentication** (e.g. `gcloud auth application-default login`) if using GCS or BigQuery.

### Steps (high level)

1. **Phase 1 — Inventory:**  
   `python3 Supplement_S2_Waveform_Extraction.py --phase inventory`  
   This scans headers and writes an inventory CSV (e.g. which subjects have PLETH, ECG, etc.).

2. **Phase 2 — Extract:**  
   `python3 Supplement_S2_Waveform_Extraction.py --phase extract --max-subjects 200`  
   (or without `--max-subjects` for full cohort). Reads PLETH segments and computes AC/DC, SNR, and related features.

3. **Phase 3 — Upload:**  
   `python3 Supplement_S2_Waveform_Extraction.py --phase upload`  
   Uploads feature tables to BigQuery (if configured).

4. Run the **Golden Subset** BigQuery queries in S1 Part G (after creating `golden_clinical_master` and `golden_spo2_paired` as in S1) to join waveform features with clinical outcomes.

Detailed step-by-step instructions (upload script to Vertex AI, install deps, run Phase 1, run first two Golden Subset queries in BigQuery) are in the project’s internal runbook; the same logic is summarized above.

---

## 4. Regression (S5)

The full-cohort and normal-lactate logistic regression models (Supplement S5) were fit in **Python** using **statsmodels**:

- **Data:** Export the table `eicu_regression_input` (S1 Part B) and, for the normal-lactate model, the table created by S1 Part F2 (`eicu_normal_lactate_regression`) to CSV.
- **Model:**  
  `logit('died ~ hidden_flag + age + C(gender) + C(ethnicity) + max_creatinine + max_bilirubin + min_platelets + max_lactate', data=df).fit(cov_type='cluster', cov_kwds={'groups': df['hospitalid']})`  
  For the normal-lactate model, omit `max_lactate` (all have lactate < 2).
- **Reference:** Coefficients and odds ratios in Supplement S5.

---

## 5. Summary checklist

| Step | Action |
|------|--------|
| 1 | Obtain PhysioNet credentialed access (MIMIC-IV, eICU, MIMIC-III, Open Oximetry as needed). |
| 2 | Link Google account to PhysioNet BigQuery for those projects. |
| 3 | Run Supplement S1 queries in BigQuery (adjust project/dataset for CREATE TABLE if needed). |
| 4 | Export regression input tables; run logistic regression (Python/statsmodels) as in S5. |
| 5 | (Optional) Download MIMIC-III Waveforms; run Supplement S2 pipeline; run S1 Part G. |

---

*For project-specific details (e.g. GCS paths, exact table names in a private dataset), see the manuscript’s data availability statement and any linked code repository.*
