## Supplementary material for "Perfusion-Dependent Melanin Bias in Pulse Oximetry and ICU Mortality Across 209 U.S. Hospitals: A Multicenter Retrospective Analysis of 52 Million Readings": Gehring_2026_PulseOx_Supplements.zip: Supplement_S7_Causal_DAG.md

# Supplement S7 — Directed Acyclic Graph (Causal Structure)

**Manuscript:** Gehring 2026. Perfusion-Dependent Melanin Bias in Pulse Oximetry and ICU Mortality Across 209 U.S. Hospitals.

This supplement describes the hypothesized causal structure linking illness severity, perfusion, melanin, pulse oximetry bias, hidden hypoxemia, and mortality. It corresponds to **Methods Section 2.8** and **Discussion Section 4.3**.

---

## Text description of the DAG

The following variables and directed edges form a simplified directed acyclic graph (DAG). The purpose is to clarify what we treat as **exposure**, **outcome**, **confounders**, and **mediators**, and why certain adjustments are used.

### Nodes

1. **Illness severity** — Unmeasured latent variable: how sick the patient is at baseline (comorbidities, acuity). Affects both perfusion and mortality.
2. **Low perfusion** — Reduced peripheral blood flow (e.g. shock, vasoconstriction). Affected by illness severity. Influences both PPG signal quality and (independently) mortality.
3. **Melanin / skin pigmentation** — Optical property of tissue. Not caused by illness in this model. Modifies the effect of low perfusion on signal quality.
4. **PPG signal quality (AC/DC)** — Strength and interpretability of the photoplethysmographic signal. Caused by perfusion and (conditionally) melanin (perfusion × melanin interaction).
5. **SpO₂ overestimation (bias)** — Device reads higher than arterial SaO₂. Caused by degraded PPG signal (and thus by low perfusion × melanin).
6. **Hidden hypoxemia** — Binary: SpO₂ ≥ 94% with SaO₂ < 88%. Caused by SpO₂ overestimation (and thus downstream of perfusion, melanin, and signal quality).
7. **Delayed escalation** — Clinical decisions (e.g. not increasing oxygen, not escalating care). Plausibly influenced by false reassurance from SpO₂ (hidden hypoxemia). Not observed in our data.
8. **In-hospital mortality** — Outcome. Caused by illness severity, low perfusion, and (plausibly) delayed escalation.

### Directed edges (arrows)

- **Illness severity → Low perfusion**  
- **Illness severity → Mortality**  
- **Low perfusion → PPG signal quality**  
- **Melanin → PPG signal quality** (effect is conditional on perfusion; no arrow from illness to melanin)  
- **Low perfusion → Mortality** (e.g. shock → death)  
- **PPG signal quality → SpO₂ overestimation (bias)**  
- **Bias → Hidden hypoxemia**  
- **Hidden hypoxemia → Delayed escalation** (plausible; not measured)  
- **Delayed escalation → Mortality**

### Implications for analysis

- **Exposure:** Hidden hypoxemia (binary, per stay).  
- **Outcome:** In-hospital mortality.  
- **Confounder:** Illness severity causes both low perfusion (hence bias/hidden hypoxemia) and mortality. We adjust for **proxies** of illness severity: age, sex, race/ethnicity, peak creatinine, peak bilirubin, nadir platelets, peak lactate.  
- **Mediators:** Creatinine and lactate are also **downstream** of hypoxia and organ failure. Adjusting for them may partially block the path from hidden hypoxemia → mortality (over-adjustment) or may capture severity (under-adjustment). We therefore report the multivariable model as **exploratory** and rely on **triangulation**: landmark analysis (first 48 h only), lactate stratification (normal lactate only), and combined landmark + normal lactate.  
- **No single analysis** fully resolves the DAG; the manuscript uses multiple complementary analyses to assess robustness.

### Minimal DAG (arrow notation)

```
Illness severity ──→ Low perfusion ──→ PPG signal quality ──→ Bias ──→ Hidden hypoxemia ──→ Delayed escalation ──→ Mortality
        │                    │                    ▲                                                                    ▲
        │                    └────────────────────┼────────────────────────────────────────────────────────────────────┘
        │                                         │
        └─────────────────────────────────────────┴──────────────────────────────────────────────────────────────────────→ Mortality
                                                  Melanin (modifies Low perfusion → PPG signal quality)
```

(Lactate and creatinine sit on paths from illness severity and from hypoxia/organ failure; they are used as severity proxies with the caveats above.)

---

*A formal diagram (e.g. drawn in DAGitty or similar software) can be generated from this description and included in a repository or extended materials.*
